# Patient symptoms and experience following COVID-19: results from a UK wide survey

**DOI:** 10.1101/2021.04.15.21255348

**Authors:** Sara C Buttery, Keir EJ Philip, Parris Williams, Andrea Fallas, Brigitte West, Andrew Cumella, Cheryl Cheung, Samantha Walker, Jennifer K Quint, Michael I Polkey, Nicholas S Hopkinson

## Abstract

**Objectives:** To investigate the experience of people who continue to be unwell after acute COVID-19, often referred to as ‘long COVID’, both in terms of their symptoms and their interactions with healthcare.

**Design:** We conducted a mixed-methods analysis (quantitative and qualitative) of responses to a survey accessed through a UK online post-COVID support and information hub between April 2020 and December 2020 about people’s experiences after having acute COVID-19.

**Participants:** Of 3290 respondents, 78% were female, median age range 45-54 years, 92.1% reported white ethnicity; 12.7% had been hospitalised. 494 respondents (16.5%) completed the survey between 4 and 8 weeks of the onset of their symptoms, 641 (21.4%) between 8 and 12 weeks and 1865 (62.1%) more than 12 weeks after.

**Results:** The ongoing symptoms most frequently reported were; breathing problems (92.1%), fatigue (83.3%), muscle weakness or joint stiffness (50.6%), sleep disturbances (46.2%), problems with mental abilities (45.9%) changes in mood, including anxiety and depression (43.1%) and cough (42.3%). Symptoms did not appear to be related to the severity of the acute illness or to the presence of pre-existing medical conditions. Analysis of free text responses revealed three main themes (1) Experience of living with COVID-19 – physical and psychological symptoms that fluctuate unpredictably; (2) Interactions with healthcare; (3) Implications for the future – their own condition, society and the healthcare system and the need for research

**Conclusion:** People living with persistent problems after the acute phase of COVID-19 report multiple and varying symptoms that are not necessarily associated with initial disease severity or the presence of pre-existing health conditions. Many have substantial unmet needs and experience barriers to accessing healthcare. Consideration of patient perspective and experiences will assist in the planning of services to address this.

**Ethical approval:** Ethical approval was granted by Imperial College Research and Integrity Team (IREC; 20IC6625).

**What we already know on this subject:**

- Many people who develop COVID-19 will go on to endure persistent symptoms past the acute phase of the disease, commonly termed long-COVID.
- Knowledge gaps exist regarding the lived experience and symptom frequency, in people with long-COVID, particularly in those who were not admitted to hospital during the acute phase of their illness.

**What this study adds:**

- The findings from this large population, many of whom were not hospitalised during the acute phase of their illness, demonstrate the varying patterns and persistence of symptoms of long-COVID, which do not appear to be associated with severity of the acute phase of the disease or pre-existing medical conditions.
- Qualitative findings revealing the patient experience of long-COVID symptoms, healthcare, and suggestions for future research and service adaptation.

## INTRODUCTION

The COVID-19 pandemic is one of the most important public health crises of recent history, with substantial impacts on human health and wellbeing, related both to the direct effects of the condition and the impact of measures to control and reduce infection (1-3). Though many people have asymptomatic infection (4) others develop COVID-19 disease. Severity of the initial phase varies dramatically between individuals, from mild flu-like symptoms to multi-organ failure and death (5). The pandemic response has largely focused on reducing hospitalisation and mortality from acute COVID-19 (6), however, a substantial proportion of people experience long term symptoms (7-13).

As a relatively new and complex phenomenon the syndromes involving persisting symptoms following acute COVID-19 are inconsistently defined (14). A recent NICE clinical guideline (15) for the management of people with symptoms following COVID-19 subcategorises this into ‘ongoing symptomatic COVID-19’ for people with persistent symptoms 4 to 12 weeks after the start of acute COVID-19; and ‘post-COVID-19 syndrome’ for those whose symptoms have not resolved 12 weeks after the onset of acute COVID-19. The different terms are also likely to be underpinned by a range of processes which are yet to be fully established, including organ damage, cognitive processing disorders, continuing inflammatory response and/or blood clotting disorders (14). In this paper we use the term “long-COVID” which appears popular among people experiencing prolonged symptoms, with some also adopting the phrase “long-haulers” (15-17).

Long-COVID is thought to affect approximately 10% of people who develop COVID-19 (15), although emerging research suggests this proportion could be far greater (18), with many experiencing symptoms for months. However, research on the patterns and persistence of longer term symptoms following acute COVID-19 (12) is still evolving, including the range and extent of symptoms and how the condition is experienced by those affected. Furthermore, much of the published data relate to people who have been hospitalised during the acute phase, rather than those who managed their acute illness in the community (7). The latter, much larger group, many of whom have never received a formal diagnosis of COVID-19 risk being overlooked by health and care services (19) and research.

To address this, we analysed responses to an online survey of people with long-COVID to explore symptom frequency, patient experience, and identify key themes to improve understanding of the condition and inform healthcare responses.

## METHODS

### Research design

We used a mixed methods approach, combining quantitative and qualitative methods, to analyse data from a UK wide survey conducted in April 2020, by the Asthma UK British Lung Foundation partnership (AUK-BLF). The survey asked respondents about the presence and duration of symptoms, the level of treatment they had required during their initial illness (e.g. at home or in hospital) and their experience of care, support and information received during and post-acute COVID-19.

The survey was accessed via the online Post-COVID hub https://www.post-covid.org.uk/. This website was developed by AUK-BLF to support individuals with ongoing symptoms following the acute illness, providing reliable information and dedicated support for their mental and physical recovery. The hub includes a dedicated nurse led helpline (WhatsApp and phone) to support people with ongoing symptoms after their acute illness, and their families. The post-COVID hub is promoted by AUK-BLF via email and social media or can be retrieved by typing ‘long-COVID support’ into a search engine. Access to the survey was via a link on the hub, but the survey itself was not publicised by AUK-BLF.

Ethical approval for this analysis was granted by Imperial College Research and Integrity Team (IREC; 20IC6625). Participants were required to give informed consent for the use of their data for this purpose at the start of the survey.

### Data analysis

Statistical analyses were performed using SPSS, version 27. Demographic data and information about pre-existing health conditions were collated and presented by groups according to the level of treatment received during the initial phase of the illness (community Vs hospitalised). Between group differences were compared using Chi-square test or independent t-tests as appropriate, with p<0.05 taken to indicate statistical significance. Where there were ordinal variables (e.g. age groups) a median of each was taken to allow independent t-tests to be calculated. For categories with very small numbers, grouping together of categories was used to account for underrepresentation. In the case of duplicate entries, the most recent was used for analysis.

### Thematic analysis

We analysed free-text responses to the survey questions: “How would you describe your experience after COVID-19?” and “Please describe what kind of follow-up support you would ideally like to receive from your healthcare providers”.

Thematic analysis was conducted by SB, PW and KP, based on the approach described by Braun and Clark (20). Following initial readings, data were entered into the NVIVO (NVIVO 1.0, QSR International) qualitative data analysis software package. We conducted an initial word frequency analysis to provide an overview of our data (see summary table and bubble chart: ***Supplement 1***). Next, we used the software to organise text extracts into common codes. From these codes, themes were developed independently by SB, PW and KP, who then came together to discuss, refine, reorganise, and agree on final themes. These were then reviewed with the co-authors, with discussion from the original data.. Quotations reported below are followed in brackets by the respondent’s gender, ethnicity, age group, any pre-existing medical condition, and time since symptom onset.

### Patient and Public involvement

Study participant validation of the themes was not possible due to the nature of data collection methods used, however the findings were discussed with and reviewed by, expert patients with long-COVID

## RESULTS

Between 28^th^ April 2020 and 15^th^ December 2020, there were 3648 responses from the AUK-BLF COVID Hub survey. We excluded 273 blank entries, and 85 duplicates, providing a final sample of 3290 responses which were included in the present analysis ***(Table 1)***. 78% of respondents were female, 92.1% reported white ethnicity and the median age range was 45-54 years. 2873 (87.3%) had remained at home during their acute illness and 417 (12.7%) had been admitted to hospital for treatment. The prevalence of asthma in our cohort was high (26.3%) in comparison to other pre-existing conditions.

**Table 1:** Demographics of all survey participants and a) those who were not hospitalised b) those who were hospitalised.

|  | All<br>n= 3290 | not hospitalised<br>n=2873 | hospitalised<br>n=417 | p-value |
| --- | --- | --- | --- | --- |
| **Gender** |  |  |  |  |
| Female | 2565 (78.0) | 2269 (79.0) | 296 (71.0) | **<0.001** |
| Male | 686 (20.9) | 568 (19.8) | 118 (28.3) |  |
| **Age** |  |  |  |  |
| 17 or under | 6 (0.2) | 4 (0.1) | 2 (0.5) | **<0.001** |
| 18-24 | 55 (1.7) | 51 (1.8) | 4 (1.0) |  |
| 24-34 | 407 (12.4) | 373 (13.0) | 34 (8.2) |  |
| 35-44 | 778 (23.6) | 702 (24.4) | 76 (18.2) |  |
| 45-54 | 1117 (34.0) | 988 (34.4) | 129 (31.0) |  |
| 55-64 | 693 (21.1) | 571 (19.9) | 122 (29.3) |  |
| 65-74 | 193 (5.9) | 153 (5.3) | 40 (9.6) |  |
| 75 or older | 40 (1.2) | 30 (1.0) | 10 (2.4) |  |
| **Ethnicity** |  |  |  |  |
| White | 3031 (92.1) | 2684 (93.4) | 379 (91.0) | 0.07 |
| Black, African, Black British or Caribbean | 41 (1.2) | 31 (1.1) | 10 (2.4) |  |
| Asian or Asian British | 91 (2.8) | 77 (2.7) | 15 (3.6) |  |
| Mixed or multiple ethnic groups | 56 (1.7) | 49 (1.7) | 7 (1.7) |  |
| **Nation** |  |  |  |  |
| England | 2742 (83.3) | 2408 (83.8) | 330 (79.1) |  |
| Scotland | 271 (8.2) | 226 (7.9) | 43 (10.3) |  |
| Wales | 155 (4.7) | 134 (4.7) | 21 (5.0) |  |
| Northern Ireland | 43 (1.3) | 30 (1.0) | 13 (3.1) |  |
| **Current work status** |  |  |  |  |
| Working full-time | 1104 (33.6) | 989 (34.4) | 111 (26.6) |  |
| Working part-time | 532 (16.2) | 483 (16.8) | 50 (12.0) |  |
| Student | 48 (1.5) | 40 (1.4) | 8 (1.9) |  |
| Retired | 304 (9.2) | 242 (8.4) | 63 (15.1) |  |
| Not working because of COVID-19 | 889 (27.0) | 744 (25.9) | 138 (33.1) |  |
| Not working because of other reasons | 267 (8.1) | 238 (8.3) | 31 (7.4) |  |
| **Total Household Income** |  |  |  |  |
| Below £20,000 | 550 (16.7) | 458 (15.9) | 93 (22.3) | **<0.001** |
| £20,000-£30,000 | 556 (16.9) | 487 (17.0) | 69 (16.5) |  |
| £30,001-£40,000 | 510 (15.5) | 341 (11.9) | 65 (15.6) |  |
| £40,001-£70,000 | 796 (24.2) | 700 (24.4) | 95 (22.8) |  |
| Over £70,000 | 496 (15.1) | 448 (15.6) | 45 (10.8) |  |
| **Pre-existing health condition** |  |  |  |  |
| Anxiety | 390 (11.9) | 335 (11.7) | 55 (13.2) | 0.37 |
| Asthma | 866 (26.3) | 723 (25.2) | 143 (34.3) | **<0.001** |
| Bronchiectasis | 32 (1.0) | 22 (0.8) | 10 (2.4) | **<0.001** |
| Cancer | 32 (1.0) | 28 (1.0) | 4 (1.0) | 0.98 |
| CVD | 58 (1.8) | 42 (1.5) | 16 (3.8) | **0.001** |
| CKD | 16 (0.5) | 11 (0.4) | 5 (1.2) | **0.025** |
| COPD | 58 (1.6) | 43 (1.5) | 14 (3.4) | **0.01** |
| Depression | 287 (8.7) | 240 (8.4) | 47 (11.3) | 0.05 |
| Diabetes | 71 (2.2) | 50 (1.7) | 21 (5.0) | **p<0.001** |
| HTN | 255 (7.8) | 202 (7.0) | 53 (12.7) | **p<0.001** |
| ILD | 2 (0.1) | 2 (0.1) | 0 (0.0) | 0.59 |
| Obesity | 218 (6.6) | 173 (6.0) | 45 (10.8) | **p<0.001** |

CVD: Cardiovascular disease; CKD: Chronic kidney disease; COPD: Chronic pulmonary disease; HTN; Hypertension; ILD: Interstitial lung disease. Data are presented as n (%). Statistical tests compare group a) and b). P values of <0.05 are for Chi<sup>2</sup> or t test as appropriate. Where numbers do not total 100% missing values are due to questions unanswered by participant.

The most frequently reported symptoms were breathing problems (92.1%), fatigue (83.3%), muscle weakness or joint stiffness (50.6%), sleep disturbances (46.2%), problems with mental abilities (45.9%), changes in mood, including anxiety and depression (43.1%) and cough (42.3%). There was no statistical difference in symptom pattern between those who had been hospitalised and the rest of the survey population ***(Figure 1 / Online Table E1)***.

**Figure 1.**
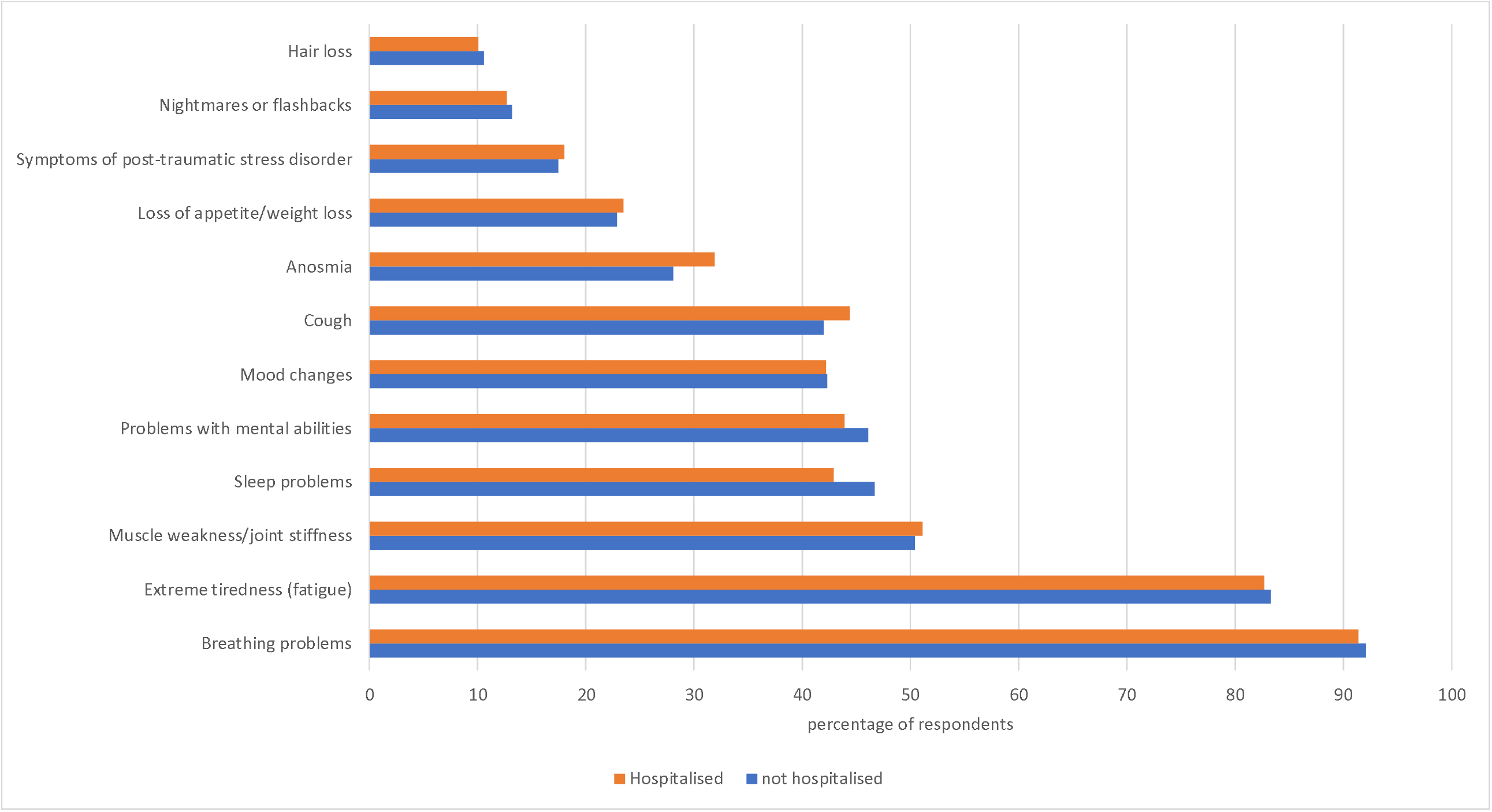
Frequency of symptoms reported by post-COVID survey respondents Graph showing percentages of individuals with long-COVID reporting each symptom. Orange bar, people hospitalised during their acute COVID-19 illness (n=417); blue bar, not-hospitalised (n=2873).

Respondents most frequently reported 5-7 co-existing symptoms (40.1%) with a mean(SD) of 5.0(2.3) ***(Figure 2)***. People with pre-existing lung disease (27.6%) were more likely to report post-COVID breathing problems compared to those without although the difference was numerically small (88.4% vs 83.7%; X^2^ 11.6, p=0.001). Changes in mood were commonly reported, however, there was no significant difference between those with or without pre-existing anxiety and/or depression (24.7% vs 22.4%) (X^2^ 1.262, p=0.261). ***(Online supplement 2)***.

**Figure 2.**
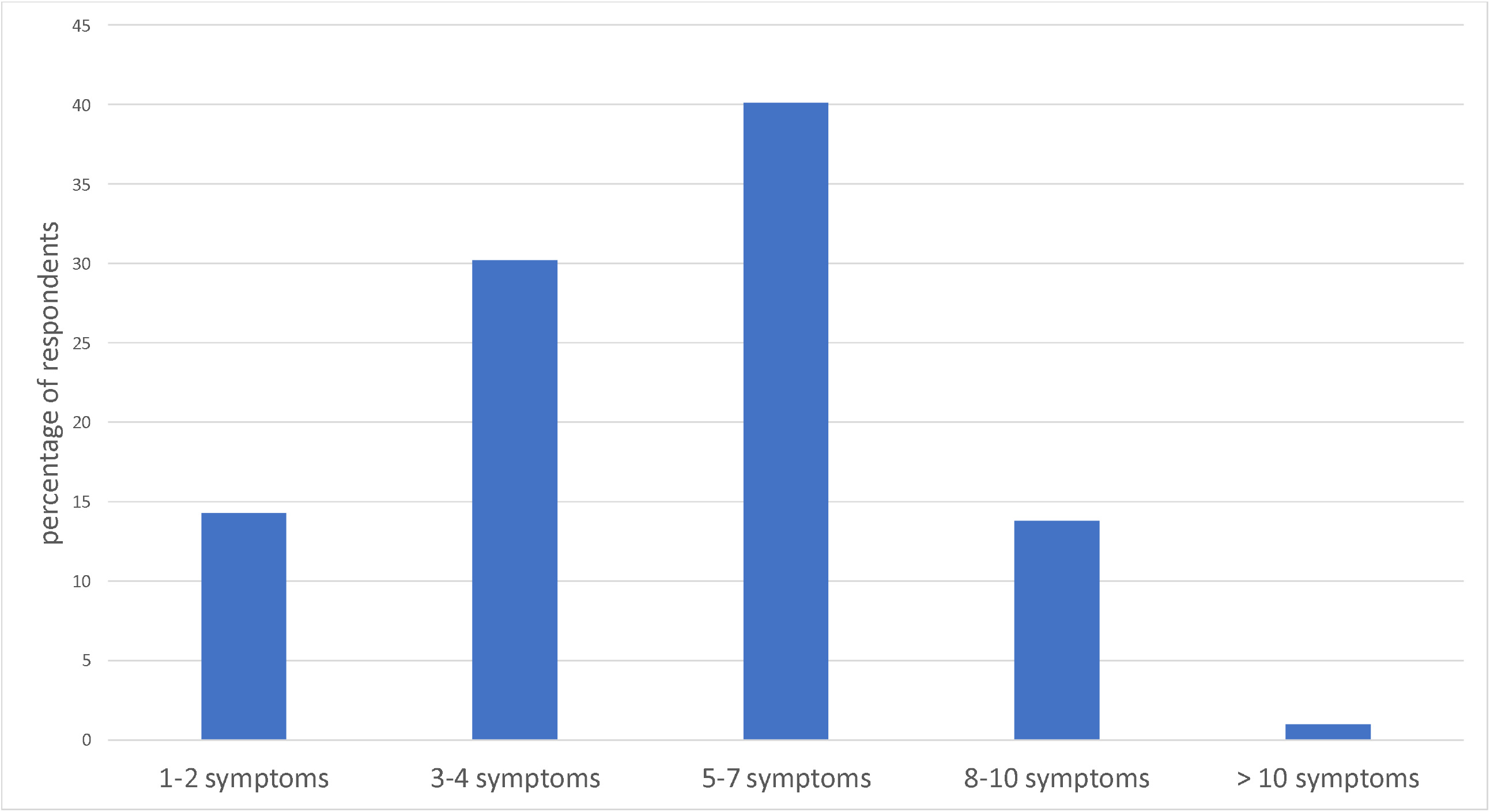
Number of symptoms reported by post-COVID survey respondents Respondents reported a mean(SD) 5.0(2.3) symptoms.

People who were hospitalised in the acute phase of their illness, compared to those who stayed at home/in the community, were more likely to be male, older, have a lower household income, and have pre-existing medical conditions including asthma, bronchiectasis, CVD, CKD, diabetes, HTN, and obesity (Table 1).

270 respondents (8.2%) completed the survey <4 weeks after the onset of symptoms, 494 respondents (15.0%) between 4 and 8 weeks of the onset of their symptoms, 642 (19.5%) between 8 and 12 weeks and 1865 (56.7%) completed the survey after 12 weeks of symptom onset. The demographics of people completing the survey at different time points were similar, with a tendency for increase in age as duration of symptoms increased ***(Table 2)***. There were some differences in the pattern of symptoms. Breathlessness and extreme tiredness or fatigue did not vary, but changes in mood, extreme tiredness or fatigue, hair loss, muscle weakness or joint stiffness, problems with mental abilities, sleep problems and symptoms of post-traumatic stress disorder were all more prevalent in those reporting after a longer interval. By contrast, symptoms of cough and loss of taste or smell were more prevalent in the earlier stages after the acute illness. Significantly more co-exiting symptoms were reported in those that reported longer on-going symptoms.

**Table 2:**
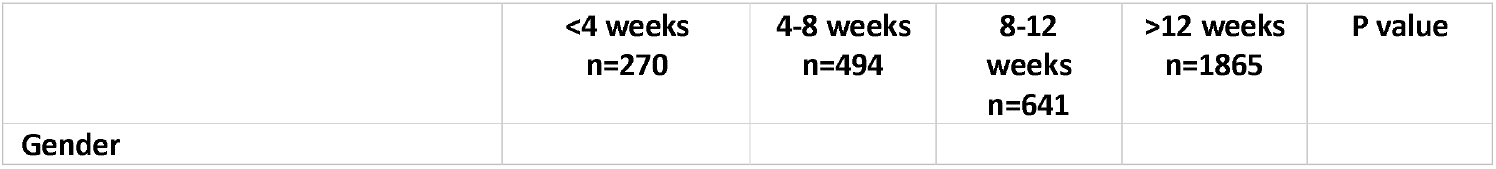
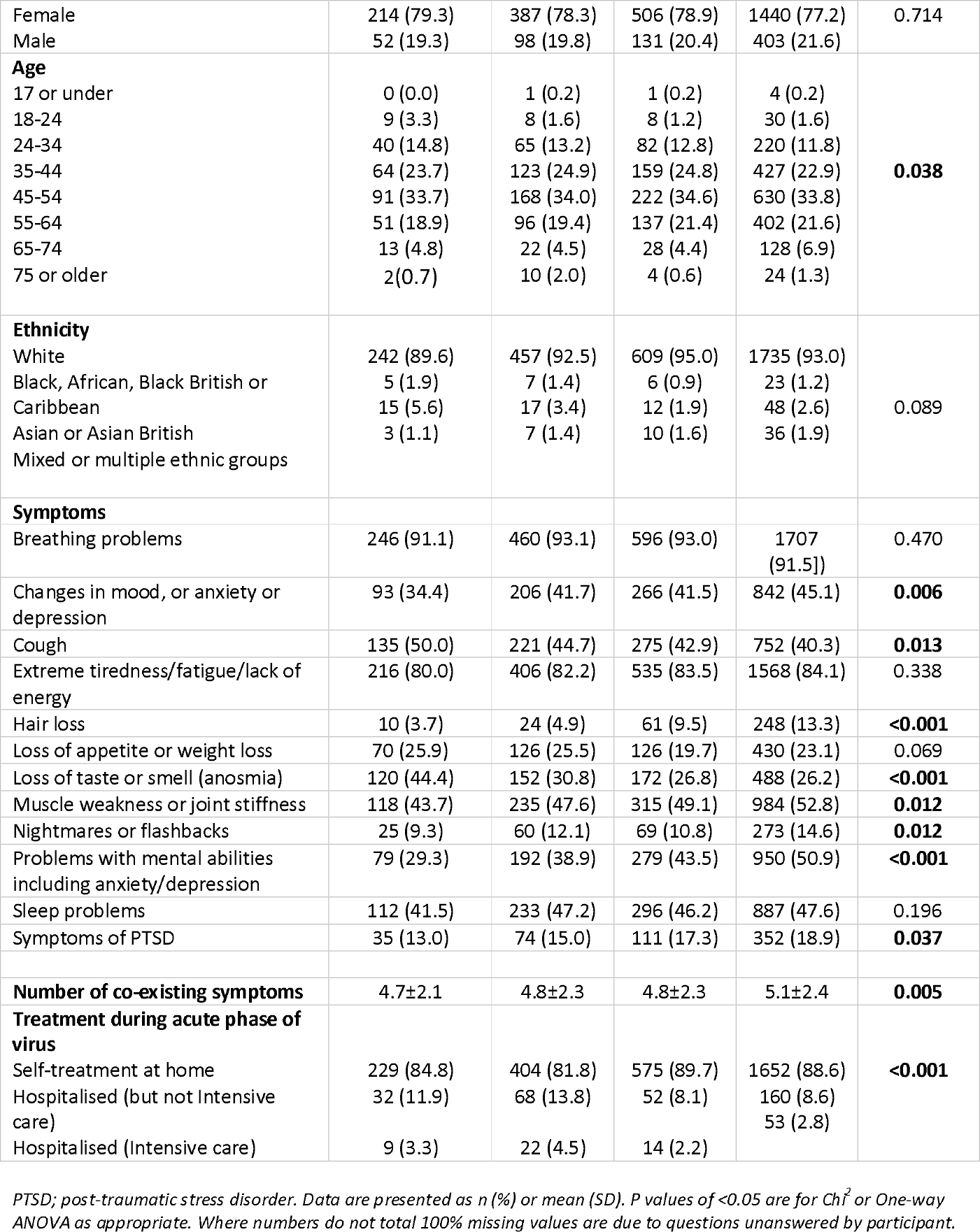
symptoms reported in those who report a) <4weeks since symptom onset; b) 4-8 weeks since symptoms onset c) 8-12 weeks since symptoms onset; d) > 12 weeks since symptoms onset.

In the 417 respondents who reported being hospitalised with COVID, 57.1% were in hospital for less than one week while 17% were in for more than 2 weeks ***(Online Table E1)***. As expected, pre-existing medical conditions were more prevalent in people who had required hospital care. Among hospitalised patients 33.6% reported not needing any sort of respiratory support during the acute phase of their illness, 50.4% required non-invasive support with their breathing (e.g. supplemental oxygen, CPAP) and 10.6% required intubation. Those who had been intubated were no more likely to report ongoing breathing difficulties, than those who had not (86.4% vs 92.6%; p=0.485). Of note, only 7% reported receiving a rehabilitation plan on discharge from hospital. 27.4% of all respondents reported that they had not spoken to their GP or nurse and only 27.9% reported being given information about things they could do to aid their recovery. The quality of communication and information received (scored on a scale of 0-10) was rated more highly in the hospitalised group in terms of clarity, usefulness, timeliness and empathy compared to those who had remained at home ***(Online Table E2)***.

In terms of importance (rated out 0 - 10), advice from healthcare professionals on managing difficulties after COVID-19 scored highest (9.0); general information on difficulties after COVID-19 (8.9); the latest medical research on difficulties after COVID-19 (8.9); and reading about and talking to other people that have difficulties after COVID-19 (8.4), were all rated highly ***(Supplement 2)***.

### Thematic analysis

Survey respondents described a range of experiences related to their condition, with many focussing on their ongoing symptoms and the condition’s impact on their daily life. Such impacts were common both in people who had been hospitalised and those who experienced the acute phase of the illness at home. Three key themes were identified: (1) Experience of living with COVID-19 – physical and psychological symptoms that fluctuate unpredictably; (2) Interactions with healthcare; (3) Implications for the future – their own condition, society and the healthcare system and the need for research ***(Figure 3)***.

**Figure 3.**
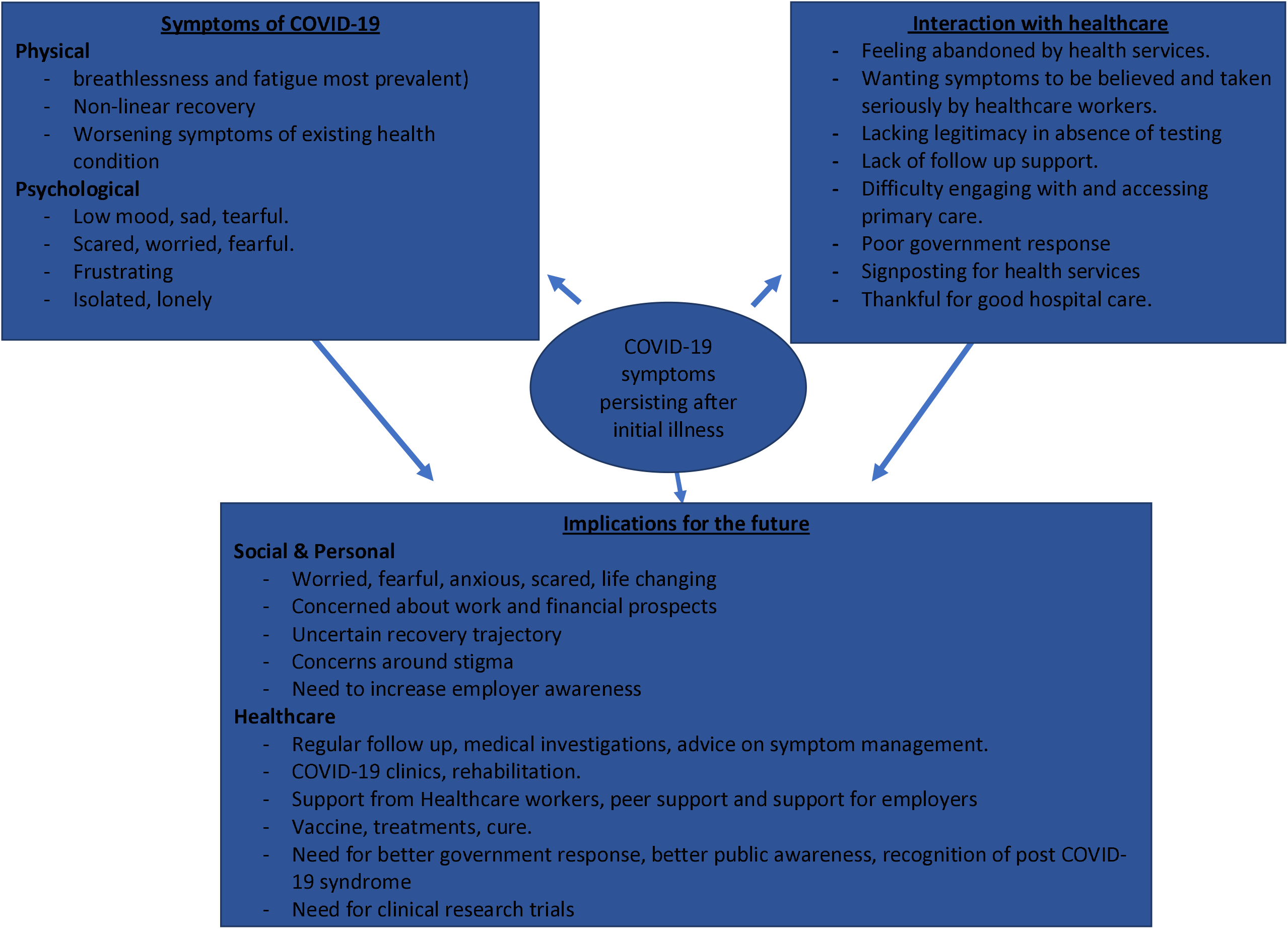
Thematic analysis of long-COVID experience.

#### Experience of living with COVID-19

Typically, respondents described multiple, often disabling symptoms causing major disruption to daily life and functioning. Both physical and psychological symptoms were common, frequently overlapping, and often exacerbated one another. Frustratingly for many, the ongoing disease course was non-linear, with recurrent and unpredictable symptomatic relapses. Participants highlighted the need to understand the long-lasting consequences of COVID-19; not just the immediate risk of death.

Many individuals reported continuing to suffer with a range of physical symptoms, the most common being breathlessness and fatigue, which mirrors our quantitative findings.

*‘Breathing is difficult on minor exertion, unable to walk upstairs without becoming short of breath, moderate exertion (walking up 4 or 5 floors) leads to a period of extreme breathlessness and exhaustion, unable to walk further’ (Male white, aged 45-54, not hospitalised, 3*.*5 months since onset of symptoms)*.

*‘Constant fatigue is a real problem’ (Female, white, 55-64, pre-existing diabetes and obesity, not hospitalised, 11 months since onset of symptoms)*.

Returning to their normal lives was almost impossible for some, and often even basic daily activities were limited, when prior to getting ill they saw themselves as fit and healthy. **(Figure 4)**

**Figure 4.**
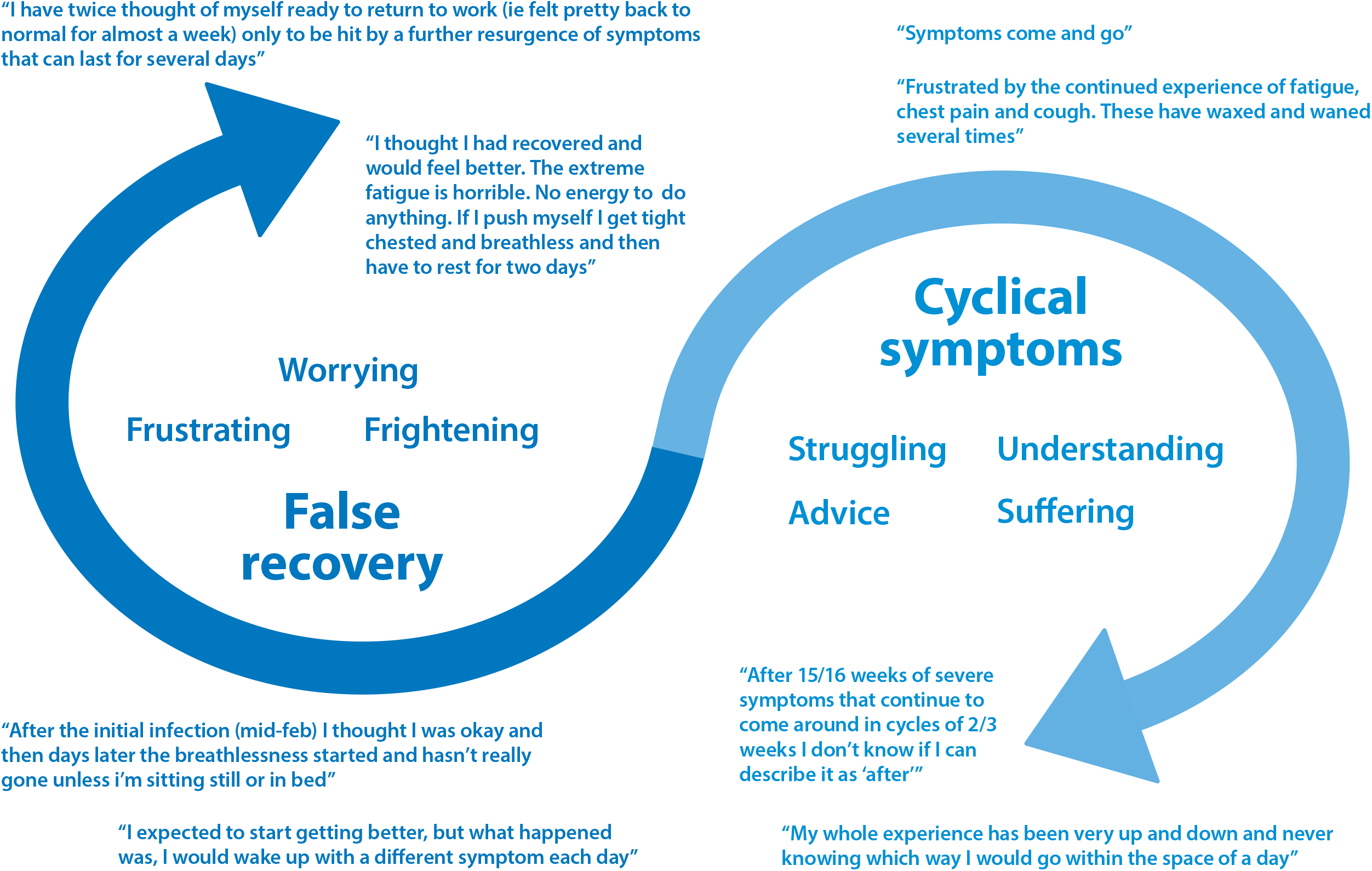
Infographic demonstrating the recovery/cyclical nature of Long-COVID symptoms from the respondent perspective.

*I just feel tired weak and sore all over. I lack the same energy I had to go about my daily life (Female, white, 35-44, pre-existing asthma, hypertension and obesity, not hospitalised, 6*.*5 months since onset of symptoms,)*.

*“I tested positive over 4 weeks ago, normally fit and healthy, now I get breathlessness really quickly. Even just walking around the house, or washing the dishes, triggers it*.*” (Female, White, 45-54, no pre-existing medical conditions, not hospitalised, 4 weeks since onset of symptoms)*

Notably, many report their disease course as non-linear, with cyclical, fluctuating symptoms and ‘false recoveries’ **(figure 4)**.

*“I feel like I’m on the mend and then have a massive set back. It is very depressing. I was doing better for the last 6 weeks, then recently had a relapse where my breathlessness is back, and my fatigue is back. Very frustrating! One step forward and three steps back” (Female, White, 25-34, no pre-existing medical condition, not hospitalised, 6*.*5 months since onset of symptoms)*

Uncertainty regarding the duration of symptoms, and completeness of recovery, was a major source of distress and was compounded by the general lack, and ambiguity, of information from healthcare providers regarding their condition **(figure 4)**.

*“Understanding what is causing breathing difficulties, e*.*g. scarring or inflammation, and evidence on how long these symptoms are likely to last. And evidence that a full recovery is likely” (Female, White, 35-44, no pre-existing medical condition, not hospitalised, 8 weeks since onset of symptoms)*

Reported psychological morbidity was pronounced with around half of respondents (43.1%) reporting persistent and debilitating anxiety. Psychological distress had multiple origins as highlighted in the themes, including physical symptoms, relapsing disease course, and barriers to care.

*“I have noticed my anxiety has increased a lot. I know it’s post-COVID symptoms but sometimes I doubt myself and think there is something else? it’s very worrying however, I can manage my symptoms but it’s more anxiety that creates more issues!” (Female, white, under 17, no pre-existing health conditions, treatment in hospital, 4 weeks since onset of symptoms)*

‘Scared’ and ‘fearful’ were repeatedly used by participants to describe their emotional status both in terms of their current position and their expectations for the future. Others described feeling lonely or isolated and low mood was common. “

*Frightening, confusing, lonely, lacking support, worried to tell people in case they don’t believe me or think I’m infectious, frustrating… it’s hard sometimes not to feel bleak due to not knowing what the long term implications are, what will happen if I get it again, are any [of] my organs damaged and going to fail in the future” (Female, White, 45-54, pre-existing anxiety disorder, not hospitalised, 12 weeks since onset of symptoms)*

### Interactions with healthcare

Among comments about accessing care, perceptions and experiences were largely dichotomised between those who felt well supported and those who did not. Thus, many participants described feeling let down or abandoned by healthcare services, consistent with almost a third (27.4%) having not spoken to a GP or nurse about their difficulties.

*“I feel very deserted and alone by health care professionals*.*” (Female, White, 55-64, pre-existing anxiety disorder, hospitalised, 7 months since onset of symptoms)*

There were many participants who described themselves as the forgotten patients of COVID-19. This cohort had experienced mild to severe symptoms but remained at home and were unable to be tested due to the lack of available tests.

*“Frustrating! No access to tests, swab and antibody, ‘til way too late, resulting in negative outcome. G. P’s refusing to refer me to specialists, insisting I was ‘fine’ when clearly not - unable to breathe/speak. Only got referral by keep pushing GP, very tiring…*.*Went to local A&E by ambulance in late June, as severe breathing problems and chest pain, 111 thought I might be having heart attack. Docs v dismissive and felt I was wasting their time. Charming!” (Female, White, 55-64, no pre-existing medical conditions, not hospitalised, 5 months since onset of symptoms)*

Experiences with the NHS 111 helpline and interacting with their GP could be negative – many felt that their primary care providers did not believe that they had long-COVID symptoms and suggested their symptoms are due to stress/ anxiety.

*“At the height of the illness in mid-March I fully expected to die. There was zero medical help, I was told to lock myself in my room and cope the best I can while struggling to breathe or move. My GP referred all COVID calls to NHS111, NHS111 took hours and if I hadn’t got through to a knowing doctor by the time I was coughing up blood, who prescribed me strong antibiotics, if COVID hadn’t got me, pneumonia would have done” (Female, mixed or multiple ethnic groups, 45-54, no pre-existing medical condition, not hospitalised, 6 months since onset of symptoms)*.

The feeling that their healthcare providers did not believe them or dismissed symptoms as being due to anxiety was frustrating.

*“Follow up for prolonged symptoms and clear self-help guidelines. To not be told it is anxiety and made to feel like it’s not real” (Female, White, 35-44, no pre-existing medical conditions, not hospitalised, 8 weeks since onset of symptoms)*

Not feeling believed by healthcare professionals was a strong theme throughout, and participants felt their symptoms were being unfairly questioned. One individual described his experience as being ‘gaslighted’ by his GP.

*“It has been an awful time and it made it worse, when I felt that I wasn’t being believed about the symptoms” (Female, White, 45-54, Previous Anxiety and Asthma diagnosis, hospitalised, 8 weeks since onset of symptoms)*.

*“I didn’t think I’d still be ill 8 months later. I feel there is very little support and my GP has ignored my symptoms. He has put it down to anxiety disorder. Which is so unfair. I know my body, and never have I experienced a burning sensation In my chest and breathing problems before” (Female, Black African, 35-44, no pre-existing medical conditions, not hospitalised, 8 months since onset of symptoms*.)

By contrast, the respondents who reported feeling well supported were largely those who had been admitted to hospital and had laboratory confirmation of COVID-19 in the acute stages of their illness. This was even more marked for those who had been admitted to an HDU/ICU, who described the care and support they received in a very positive light.

*“I’ve been quite happy with the treatment I’ve received in my local hospital it’s been reassuring breathing space keeping in touch” (Female, White, 65-74, pre-existing asthma, High blood pressure and obesity, hospitalised, unknown symptom duration)*.

### Implications for the future

A third key theme centred on individuals’ future prospects and uncertainty regarding their symptoms, disease course and hopes for recovery to normal work and social life; societal and health professionals understanding of their illness and structures of care; and need for research and better treatments.

Participants described their experience with COVID-19 as ‘life-changing’ with implications in many aspects of their life. The ongoing daily battle or ‘fight’ was continually described as exhausting, with individuals despairing about their prospects in the future.

*“Frustrating and life changing. Feel very isolated from normal life and that my life has come to a shuddering halt” (Male, White, England, South-East, 45-54, no pre-existing medical conditions, not hospitalised, 6 months since onset of symptoms)*.

Responses frequently addressed concerns related to their symptoms and employment situation. Many had not been able to return to their full duties at work, others not at all.

*“can’t go back to work as I still feel so unwell and my work (I am a teacher) are trying to push me into going back as I’m out of the 14-day infectious period and they are short staffed*.*” (Female, white, 45-54, pre-existing diagnosis of bronchiectasis, not hospitalised, 4 weeks since onset of symptoms)*.

This caused significant financial stress, without a firm diagnosis to allow them to be signed off work or any certainty as to when things may get better. Increased need for extra help and care since contracting the virus was common. For many this came from family members causing extra stress on loved ones and their relationships. For those requiring paid external care, the lack of funding available for this was noted.

*“I feel there is not much help for survivors in terms of emotional and financial support. One is left to figure out how best to manage on their own… because you do not have the financial support especially as a single parent you have to force and drag yourself to work in order to pay bills. Instead of concentrating on the healing process you are left with no choice but to work*.*” (Female, Black, African, Black British or Caribbean, 55-64, pre-existing diagnosis of Hypertension and obesity, not hospitalised, 8 weeks since onset of symptoms)*

Participants expressed their hopes for the future in terms of increasing knowledge and awareness as well as access to further support interventions. This was needed especially for those who had not been admitted to hospital but were still experiencing longer lasting symptoms of the virus. In this context, more medical investigations including chest x-rays, blood test analysis and antibody testing were requested to identify and confirm disease status.

*“I would really like a thorough examination, (chest x-ray for example) to determine precisely what the problems are and how best they can be resolved*.*” (Male, White, 75 or older, no pre-existing medical conditions, hospitalised, 4 months since onset of symptoms)*.

*“More access to medical checks for reassurance and an antibody test as my case was unconfirmed due to lack of testing at the time*.*” (Female, White, 35-44, Anxiety disorder, not hospitalised, 3 months since onset of symptoms)*.

Others considered the availability of long-COVID clinics to be of great value in providing ongoing support, advice and treatment for recovery from the virus.

*“Clinics set up with multi-disciplinary teams to address all aspects of COVID recovery would be the best way forward for us all” (Female, White, 45-54, ME diagnosis, not hospitalised, 4 months since onset of symptoms)*

As well as a desire for recovery and return to ‘normal life’ there was also a need for more practical advice of how persistent symptoms could be managed. Suggestions such as increased rehabilitation services, breathing control exercises and energy conservation or pacing strategies were frequently mentioned, implying participants were anticipating or had experienced a very slow or incomplete recovery.

*“A programme of exercises starting from very simple movements and building up to more strenuous that can be used to rebuild strength & stamina gradually as fatigue & general malaise allow, taking into consideration the frequent relapses I have experienced every time I begin to improve a little!” (Female, white, 65-74, pre-existing asthma, not hospitalised, 3 months since onset of symptoms*.)

*“More information on how to cope with breathlessness. Techniques of breathing to help symptoms” (Female, White, 45-54, pre-existing anxiety, not hospitalised, 11 weeks since onset of symptoms)*.

Hopes for future research into the longer-term effects of COVID-19 was repeatedly mentioned in relation to vaccination, new treatments and a cure but also more generally to increase the knowledge base and understanding of the public, NHS and government.

*“That there is government advice for people with long-COVID. Recognition, guidance and support is not there” (Female, White, 45-54, no pre-existing medical conditions, not hospitalised, 4 months since onset of symptoms)*.

*“There needs to be more information and research looking into individuals experiencing pro longed mild/moderate (I*.*e*., *more than 4 weeks) symptoms during recovery for COVID” (female, White, 25-34, no pre-existing medical conditions, not hospitalised, 6 weeks since onset of symptoms)*.

## DISCUSSION

The results from this large survey of people living with Long-COVID demonstrate the breadth and scale of the burden they experience in terms of: multiple, varying physical and psychological symptoms that are not necessarily related to initial illness severity or pre-existing health conditions, interactions with healthcare which are often unsatisfactory in relation to both treatment that is available and the perceived legitimacy of their condition, and uncertainty about their future. Evidently, major barriers to accessing healthcare and follow-up support exist and are experienced especially by people who managed the acute phase of their illness at home and did not have laboratory confirmation of COVID-19. By contrast, those treated in hospital, who had confirmation of their diagnosis, reported fewer barriers to care. These substantial unmet care needs call for holistic approaches to support recovery.

### Significance of findings

Our findings demonstrate the varying patterns and persistence of symptoms of Long-COVID, which do not necessarily appear to be associated with severity of the acute phase of the virus and are in keeping with those presented by other recent papers(7-12). The most common symptoms reported in this study of people with long-COVID were breathlessness and fatigue. A European study of 2113 people recruited online, also found that fatigue and dyspnoea were the most prevalent symptoms post-acute COVID-19 with a median of six symptoms per person persisting (21). We found that rates of reporting post-COVID breathing problems differed by only a small amount between those who did or did not have pre-existing lung disease, consistent with a previous small study (12). Interestingly, the Zoe COVID Symptom Study has identified asthma as the only pre-existing long term condition associated with an increased risk of developing Long-COVID (22).

The variation in symptoms described supports the concept of Long-COVID comprising multiple different, often overlapping, and coexisting syndromes (14). Having taken a mixed methods approach, our findings illustrate not only the symptoms, but also how they are impacting people’s lives, the inclusion of which in guidelines and services is vital (23). The themes that emerged from analysis of responses to this survey support those of a study, using interviews and focus groups to study lived experiences of long-COVID in 114 participants (19), which identified comparable themes including access to healthcare services, concepts of improvements for the future, and supported the need to establish quality principles for a long-COVID service. Our work strengthens these findings, building the evidence base around the lived experience of long-COVID in a larger cohort.

Many of our respondents reported difficulties when interacting with primary care services regarding their symptoms. Funding for long-COVID services from NHS England was launched in November 2020 (24). However, access to these services requires referral from a General Practitioner (GP) and participant responses highlight that a greater understanding of how long-COVID can be recognised and distinguished from other possible causes of symptoms is needed. Acknowledging the legitimacy of patient symptoms and diagnosis, especially in the absence of testing at the time of their initial illness is important. NICE recently published best practice guidance on identifying and assessing those with symptoms post COVID-19 and recommendations for management and follow-up (16). Further research efforts and initiatives are responding to the impact on individuals’ lives caused by the pandemic (25) such as the NHS hub that has been set up for sufferers (26). Patients’ knowledge must be included in planning long-COVID care including the development of public understanding and perception, healthcare management strategies and how research is prioritised (27). The potential of providing such a service has been explored, with novel virtual models being supported (28-30) and its role in supporting physical and cognitive recovery (31), utilising multi-disciplinary input to tackle a multitude of needs has been evaluated (31). However, that study only looked at a cohort of patients that had been admitted to hospital and thus far has not been explored in those patients who remained at home but continue to experience ongoing symptoms.

### Methodological issues

This is one of the largest studies to date exploring the lived experience of people with long-COVID, and the inclusion of a large number of people who managed their acute illness at home provides important insight into this largely unseen group. Some limitations should be noted. Firstly, although a large sample, it is unclear how representative it is of all people with long-COVID, as our sample has high proportions of women and white people. However, emerging research suggests that females may be at an increased risk of developing long-COVID (14). Although the prevalence of asthma was 26% in our cohort, this was similar to that seen in the ISARIC dataset; 21% for 16 to 49 year olds(32). As the population are self-selecting, based on a decision to visit an online post-COVID resource,, caution should be applied to inferences regarding the factors that may predict the development of long-COVID in any one individual.

A degree of sampling bias towards those with higher levels of digital literacy, and individuals with more severe ongoing symptoms may have occurred, as such individuals could plausibly be more likely to be seeking online sources of support such as through the post-COVID hub.

## Conclusion

The three themes we identified each require specific attention to improve the understanding and management of long-COVID. 1) Symptoms – how to relieve them and better understand them; 2) Healthcare – how to make it accessible for all suffering with the lasting effects of COVID-19, not just those admitted to hospital, and in particular those who did not have a COVID-19 test at the time of their initial illness 3) Uncertainty - can be reduced by communicating existing knowledge more effectively and undertaking research to improve understanding of mechanisms and prognosis.

The COVID-19 pandemic reaches far beyond the acute phase of the illness. People with Long-COVID have substantial unmet care needs, and although services are being developed to address these needs, many people experience prohibitive barriers, including feeling that their condition is not being taken seriously by healthcare providers.

## Data Availability

Data may be made available on reasonable request.

## Contributors

SB and KEJP carried out the quantitative analysis. SB, KEJP and PW conducted the thematic analysis. SB wrote the first draft to which all authors contributed. The survey was developed by AUK-BLF partnership (AF, AC, SW, BW). All authors have reviewed and approved the final version. The corresponding author attests that all listed authors meet authorship criteria and that no others meeting the criteria have been omitted

## Ethics approval

Ethical approval was granted by the Imperial College Research Governance and Integrity Team (RGIT) (ICREC Ref: 20IC6625). All survey respondents consented to the use of their responses for analysis and publication.

## Transparency declaration

NSH, the manuscript’s guarantor affirms that the manuscript is an honest, accurate, and transparent account of the study being reported; that no important aspects of the study have been omitted; and that any discrepancies from the study as planned (and, if relevant, have been explained.

## Data Sharing

Data may be made available on reasonable request.

## Dissemination to participants and related patient and public communities

Dissemination to individual participants will not be possible but results will be shared with AUK-BLF for wider dissemination.

