## Supplement 1 for "Patient symptoms and experience following COVID-19: results from a UK wide survey"

| **Word** | **Length** | **Count** | **Weighted Percentage** | **Similar Words** |
| --- | --- | --- | --- | --- |
| feel | 4 | 2063 | 1.54% | feel, feeling, feelings, feels |
| covids | 6 | 1809 | 1.35% | 'covid, covid, covid', covids |
| symptoms | 8 | 1757 | 1.31% | symptom, symptoms, symptoms' |
| getting | 7 | 1508 | 1.13% | 'get, get, gets, getting |
| weeks | 5 | 1314 | 0.98% | week, weekly, weeks |
| helps | 5 | 1199 | 0.90% | help, help', helped, helpful, helping, helps |
| likely | 6 | 1181 | 0.88% | like, liked, likely, likes |
| breaths | 7 | 1161 | 0.87% | breath, breathe, breathe', breathed, breathes, breathing, breaths |
| tests | 5 | 1105 | 0.83% | test, tested, testing, tests |
| times | 5 | 1023 | 0.76% | time, time', timely, times |
| know | 4 | 1014 | 0.76% | know, knowing, knows |
| still | 5 | 1008 | 0.75% | still |
| days | 4 | 978 | 0.73% | 'day, day, day', days |
| long | 4 | 975 | 0.73% | 'long, long, longed |
| support | 7 | 938 | 0.70% | support, supported, supporters, supporting, supportive |
| working | 7 | 937 | 0.70% | work, worked, working, works |
| now | 3 | 890 | 0.67% | now |
| chest | 5 | 783 | 0.59% | chest, chested, chests |
| people | 6 | 739 | 0.55% | people, people', peoples |
| just | 4 | 733 | 0.55% | 'just, just |
| back | 4 | 720 | 0.54% | back, back', backs |
| needs | 5 | 638 | 0.48% | need, needed, needing, needs |
| illnesses | 9 | 633 | 0.47% | ill, illness, illnesses |
| better | 6 | 603 | 0.45% | better, better', better’ |
| pains | 5 | 592 | 0.44% | pain, painful, pains |
| informing | 9 | 588 | 0.44% | inform, informant, information, informative, informed, informing, informs |
| hospitals | 9 | 558 | 0.42% | hospital, hospitalization, hospitalized, hospitals |
| lungs | 5 | 552 | 0.41% | lung, lungs |
| months | 6 | 544 | 0.41% | month, monthly, months, months' |
| posting | 7 | 502 | 0.38% | post, posted, posting |
| recovery | 8 | 499 | 0.37% | 'recovery', recoveries, recovery, recovery' |
| one | 3 | 483 | 0.36% | one, ones |
| also | 4 | 464 | 0.35% | also |
| breathlessness | 14 | 462 | 0.35% | breathless, breathlessness |
| doctor | 6 | 460 | 0.34% | doctor, doctors |
| take | 4 | 457 | 0.34% | take, takes, taking |
| worrying | 8 | 457 | 0.34% | worried, worries, worry, worrying, worryingly |
| fatigue | 7 | 455 | 0.34% | fatigue, fatigue', fatigued, fatigues |
| normally | 8 | 455 | 0.34% | 'normal, 'normal', normal, normality, normally |
| medics | 6 | 453 | 0.34% | medic, medical, medically, medicate, medicated, medicating, medication, medications, medics |
| advice | 6 | 443 | 0.33% | advice, advices |
| health | 6 | 442 | 0.33% | health |
| going | 5 | 442 | 0.33% | going, going' |
| think | 5 | 436 | 0.33% | think, thinking, thinks |
| recovering | 10 | 429 | 0.32% | 'recovered', recover, recovered, recovering, recovers |
| understanding | 13 | 428 | 0.32% | understand, understandable, understandably, understanding, understands |
| life | 4 | 427 | 0.32% | life, life' |
| even | 4 | 421 | 0.31% | even, evening, evenings |
| really | 6 | 412 | 0.31% | really |
| never | 5 | 409 | 0.31% | never |
| able | 4 | 406 | 0.30% | able |
| much | 4 | 404 | 0.30% | much |
| coughs | 6 | 400 | 0.30% | 'cough, cough, coughed, coughing, coughs |
| frustrating | 11 | 395 | 0.30% | frustrated, frustrating, frustratingly, frustration, frustrations |
| homes | 5 | 389 | 0.29% | home, homes |
| virus | 5 | 380 | 0.28% | virus, viruses |
| felt | 4 | 379 | 0.28% | felt |
| follow | 6 | 374 | 0.28% | follow, followed, follower, following, follows |
| walk | 4 | 373 | 0.28% | walk, walked, walking, walks |
| asthma | 6 | 370 | 0.28% | asthma, asthma' |
| exercising | 10 | 362 | 0.27% | exercise, exercised, exerciser, exercises, exercising |
| problems | 8 | 357 | 0.27% | problem, problems |
| terms | 5 | 355 | 0.27% | term, terms |
| struggling | 10 | 354 | 0.26% | struggle, struggled, struggles, struggling |
| suffering | 9 | 352 | 0.26% | suffer, suffered, sufferer, sufferers, suffering, suffers |
| seem | 4 | 345 | 0.26% | seem, seemed, seemingly, seems |
| want | 4 | 340 | 0.25% | want, wanted, wanting, wants |
| caring | 6 | 337 | 0.25% | care, cared, careful, cares, caring |
| seeing | 6 | 331 | 0.25% | see, seeing |
| itâ | 3 | 328 | 0.25% | itâ |
| experiences | 11 | 320 | 0.24% | experience, experiences, experiences', experiment |
| trying | 6 | 319 | 0.24% | 'try, tried, try, tryed, trying |
| told | 4 | 309 | 0.23% | told |
| using | 5 | 309 | 0.23% | use, used, useful, usefully, using |
| things | 6 | 309 | 0.23% | thing, things |
| starts | 6 | 308 | 0.23% | start, started, starting, starts |
| worse | 5 | 304 | 0.23% | worse |
| activity | 8 | 300 | 0.22% | activate, activation, active, actively, activities, activities', activity |
| make | 4 | 297 | 0.22% | make, makes, making |
| left | 4 | 292 | 0.22% | left |
| anxiety | 7 | 291 | 0.22% | anxiety, anxiety' |
| since | 5 | 289 | 0.22% | since |
| issues | 6 | 284 | 0.21% | issue, issued, issues |
| frightening | 11 | 284 | 0.21% | frightened, frightening, frightens |
| effects | 7 | 283 | 0.21% | effect, effected, effecting, effective, effectively, effectiveness, effects |
| last | 4 | 280 | 0.21% | last, lasted, lasting, lastly, lasts |
| return | 6 | 279 | 0.21% | return, returned, returning, returns |
| calls | 5 | 274 | 0.20% | call, called, calling, calls |
| experiencing | 12 | 271 | 0.20% | experienced, experiencing |
| lot | 3 | 268 | 0.20% | lot, lots |
| exhaustion | 10 | 268 | 0.20% | exhaust, exhausted, exhausting, exhaustion, exhausts |
| donâ | 4 | 264 | 0.20% | donâ |
| manage | 6 | 260 | 0.19% | manage, managed, management, manager, managers, managing |
| improve | 7 | 256 | 0.19% | improve, improved, improvement, improvements, improves, improving |
| difficult | 9 | 254 | 0.19% | difficult |
| checks | 6 | 254 | 0.19% | check, checked, checking, checks |
| blood | 5 | 252 | 0.19% | blood, blooded, bloods |
| find | 4 | 249 | 0.19% | find, finding, findings, finds |
| well | 4 | 249 | 0.19% | well, well', wellness |
| thank | 5 | 248 | 0.19% | thank, thankful, thankfully, thanks |
| fits | 4 | 245 | 0.18% | fit, fitness, fits, fitting |
| due | 3 | 245 | 0.18% | due |
| shortness | 9 | 237 | 0.18% | short, shortly, shortness |
| antibody | 8 | 237 | 0.18% | antibodies, antibody |
| isolation | 9 | 237 | 0.18% | isolate, isolated, isolating, isolation |
| scary | 5 | 236 | 0.18% | scary |
| coming | 6 | 235 | 0.18% | 'coming, come, comes, coming |
| physical | 8 | 235 | 0.18% | physical, physically |
| severity | 8 | 233 | 0.17% | sever, several, severe, severely, severity, severly |
| got | 3 | 232 | 0.17% | got |
| anxious | 7 | 232 | 0.17% | 'anxious', anxious, anxious' |
| first | 5 | 232 | 0.17% | first, firstly |
| lack | 4 | 230 | 0.17% | lack, lacking |
| difficulties | 12 | 230 | 0.17% | difficulties, difficulty |
| bad | 3 | 228 | 0.17% | bad, badly |
| alone | 5 | 227 | 0.17% | alone |
| hardly | 6 | 224 | 0.17% | hard, hardly |
| nhs | 3 | 224 | 0.17% | nhs |
| little | 6 | 220 | 0.16% | little |
| ongoing | 7 | 218 | 0.16% | ongoing |
| body | 4 | 216 | 0.16% | bodies, body, bodys |
| good | 4 | 215 | 0.16% | good, goodness, goods |
| anything | 8 | 215 | 0.16% | anything |
| heart | 5 | 214 | 0.16% | heart |
| many | 4 | 214 | 0.16% | many |
| infections | 10 | 213 | 0.16% | infect, infected, infection, infections, infective |
| may | 3 | 209 | 0.16% | may |
| scares | 6 | 208 | 0.16% | scare, scared, scares, scaring |
| without | 7 | 208 | 0.16% | without |
| tiring | 6 | 207 | 0.15% | tire, tired, tires, tiring |
| rests | 5 | 206 | 0.15% | 'rest', rest, rested, resting, rests |
| phoning | 7 | 206 | 0.15% | phone, phoned, phones, phoning |
| treatments | 10 | 205 | 0.15% | 'treatment', treatment, treatments |
| patients | 8 | 203 | 0.15% | patient, patients |
| fears | 5 | 202 | 0.15% | fear, feared, fearful, fearfully, fearing, fears |
| extremely | 9 | 200 | 0.15% | extreme, extremely |
| inhalers | 8 | 199 | 0.15% | inhalation, inhalations, inhale, inhaler, inhalers, inhaling |
| damage | 6 | 199 | 0.15% | damage, damage', damaged, damages, damaging |
| believe | 7 | 197 | 0.15% | believe, believed, believer, believes, believing |
| professionals | 13 | 196 | 0.15% | professional, professionals |
| mental | 6 | 194 | 0.15% | mental, mentally |
| available | 9 | 193 | 0.14% | avail, availability, available |
| unable | 6 | 193 | 0.14% | unable |
| contact | 7 | 190 | 0.14% | contact, contacted, contacting, contacts |
| thought | 7 | 190 | 0.14% | thought, thoughts |
| continuous | 10 | 190 | 0.14% | continual, continually, continuation, continue, continued, continues, continuing, continuity, continuous, continuously |
| mildly | 6 | 190 | 0.14% | 'mild', mild, mildly, mildly' |
| tightness | 9 | 188 | 0.14% | tight, tightness |
| ends | 4 | 188 | 0.14% | end, ended, ending, ends |
| positive | 8 | 187 | 0.14% | position, positive, positive', positively |
| two | 3 | 186 | 0.14% | two |
| happening | 9 | 180 | 0.13% | happen, happened, happening, happens |
| concerns | 8 | 179 | 0.13% | concern, concerned, concerning, concerns |
| living | 6 | 178 | 0.13% | live, lived, lives, living |
| every | 5 | 178 | 0.13% | 'every, every |
| ray | 3 | 176 | 0.13% | ray, rays |
| hoping | 6 | 175 | 0.13% | hope, hoped, hopeful, hopefully, hoping |
| whether | 7 | 174 | 0.13% | whether |
| clearly | 7 | 172 | 0.13% | clear, cleared, clearing, clearly, clears |
| access | 6 | 171 | 0.13% | access, accessed, accessible, accessing |
| changing | 8 | 171 | 0.13% | change, changed, changes, changing |
| sleeping | 8 | 169 | 0.13% | sleep, sleeping |
| differing | 9 | 168 | 0.13% | differ, difference, differences, different, differently, differing |
| expecting | 9 | 168 | 0.13% | 'expect', expect, expectancy, expectation, expectations, expected, expecting, expects |
| research | 8 | 168 | 0.13% | research, researched, researchers, researching |
| affects | 7 | 167 | 0.12% | affect, affected, affecting, affection, affects |
| made | 4 | 167 | 0.12% | made |
| given | 5 | 166 | 0.12% | given |
| personally | 10 | 165 | 0.12% | 'personal, person, personal, personality, personalized, personally |
| enough | 6 | 165 | 0.12% | enough |
| ever | 4 | 163 | 0.12% | ever |
| others | 6 | 163 | 0.12% | others |
| nursing | 7 | 163 | 0.12% | nurse, nursed, nurses, nursing |
| group | 5 | 163 | 0.12% | group, groups |
| canâ | 4 | 162 | 0.12% | canâ |
| cause | 5 | 161 | 0.12% | cause, caused, causes, causing |
| family | 6 | 159 | 0.12% | families, family |
| later | 5 | 159 | 0.12% | later |
| though | 6 | 159 | 0.12% | though |
| negative | 8 | 158 | 0.12% | negative, negatively, negatives, negativity |
| gps | 3 | 158 | 0.12% | gps |
| unwell | 6 | 158 | 0.12% | unwell, unwell' |
| year | 4 | 157 | 0.12% | year, years |
| surely | 6 | 156 | 0.12% | sure, surely |
| way | 3 | 153 | 0.11% | way, ways |
| etc | 3 | 153 | 0.11% | etc |
| facing | 6 | 152 | 0.11% | face, faced, faces, facing |
| relapse | 7 | 148 | 0.11% | relapse, relapsed, relapses, relapsing |
| full | 4 | 147 | 0.11% | full, fullness |
| keeps | 5 | 146 | 0.11% | keep, keeping, keeps |
| talk | 4 | 146 | 0.11% | talk, talked, talking |
| looks | 5 | 146 | 0.11% | 'looked, look, looked, looking, looks |
| respiratory | 11 | 146 | 0.11% | respiratory |
| cases | 5 | 145 | 0.11% | case, cases |
| 111 | 3 | 144 | 0.11% | 111 |
| hours | 5 | 144 | 0.11% | hour, hourly, hours |
| seriously | 9 | 144 | 0.11% | serious, seriously, seriousness |
| generally | 9 | 144 | 0.11% | general, generally |
| asked | 5 | 143 | 0.11% | ask, asked, asking, asks |
| depression | 10 | 142 | 0.11% | depressant, depressants, depressed, depressing, depression, depressive |
| lonely | 6 | 141 | 0.11% | lonely |
| new | 3 | 140 | 0.10% | new |
| found | 5 | 139 | 0.10% | found, founded |
| awareness | 9 | 139 | 0.10% | 'aware', aware, awareness |
| antibiotics | 11 | 138 | 0.10% | antibiotic, antibiotics |
| constant | 8 | 137 | 0.10% | constant, constantly |
| taken | 5 | 137 | 0.10% | taken |
| completely | 10 | 134 | 0.10% | complete, completed, completely, completing |
| appointment | 11 | 134 | 0.10% | appointment, appointments |
| sickness | 8 | 134 | 0.10% | sick, sickness |
| confusing | 9 | 133 | 0.10% | confused, confusing, confusion |
| march | 5 | 132 | 0.10% | march |
| initiative | 10 | 131 | 0.10% | initial, initially, initiated, initiative |
| nothing | 7 | 130 | 0.10% | nothing |
| someone | 7 | 130 | 0.10% | someone |
| clinics | 7 | 130 | 0.10% | clinic, clinical, clinically, clinics |
| answers | 7 | 129 | 0.10% | answer, answered, answering, answers |
| employment | 10 | 129 | 0.10% | employed, employer, employers, employment |
| done | 4 | 129 | 0.10% | done |
| quite | 5 | 128 | 0.10% | quit, quite |
| around | 6 | 127 | 0.09% | around |
| levels | 6 | 127 | 0.09% | level, levels |
| yet | 3 | 126 | 0.09% | 'yet', yet |
| waiting | 7 | 126 | 0.09% | wait, waited, waiting |
| bed | 3 | 126 | 0.09% | bed, bedding, beds |
| putting | 7 | 126 | 0.09% | put, puts, putting |
| viral | 5 | 125 | 0.09% | viral |
| possibly | 8 | 125 | 0.09% | possibilities, possibility, possible, possibly |
| runs | 4 | 123 | 0.09% | run, running, runs |
| energy | 6 | 122 | 0.09% | energy |
| speak | 5 | 121 | 0.09% | speak, speaking, speaks |
| went | 4 | 121 | 0.09% | went |
| services | 8 | 120 | 0.09% | 'service', service, services |
| might | 5 | 119 | 0.09% | might |
| dying | 5 | 118 | 0.09% | die, died, dieing, dying |
| deal | 4 | 117 | 0.09% | deal, dealing, deals |
| although | 8 | 117 | 0.09% | although |
| especially | 10 | 117 | 0.09% | especially |
| scan | 4 | 116 | 0.09% | scan, scanned, scans |
| offered | 7 | 116 | 0.09% | offer, offered, offering, offers |
| awful | 5 | 116 | 0.09% | awful |
| didnâ | 5 | 116 | 0.09% | didnâ |
| condition | 9 | 115 | 0.09% | condition, conditioning, conditions |
| healthy | 7 | 115 | 0.09% | healthy |
| giving | 6 | 114 | 0.09% | give, gives, giving |
| self | 4 | 114 | 0.09% | 'self, self |
| nights | 6 | 113 | 0.08% | night, nightly, nights |
| regular | 7 | 113 | 0.08% | regular, regularly |
| governments | 11 | 111 | 0.08% | governement, government, governments |
| debilitating | 12 | 111 | 0.08% | debilitated, debilitating, debilitation |
| early | 5 | 110 | 0.08% | early |
| fevers | 6 | 110 | 0.08% | fever, fevers |
| treating | 8 | 109 | 0.08% | treat, treated, treating |
| right | 5 | 109 | 0.08% | right, rightly, rights |
| coping | 6 | 108 | 0.08% | cope, coped, coping |
| daily | 5 | 108 | 0.08% | daily |
| longer | 6 | 107 | 0.08% | longer |
| reassurance | 11 | 106 | 0.08% | reassurance, reassure, reassured, reassuring |
| results | 7 | 106 | 0.08% | result, resulted, resulting, results |
| become | 6 | 106 | 0.08% | become, becomes, becoming |
| properly | 8 | 106 | 0.08% | 'proper', 'properly, proper, properly |
| stays | 5 | 106 | 0.08% | 'stay, stay, stayed, staying, stays |
| highly | 6 | 105 | 0.08% | high, highly |
| leave | 5 | 105 | 0.08% | leave, leaves, leaving |
| impact | 6 | 104 | 0.08% | impact, impacted, impactful, impacting, impacts |
| relatives | 9 | 104 | 0.08% | relate, related, relates, relating, relation, relative, relatively, relatives |
| listens | 7 | 104 | 0.08% | listen, listened, listening, listens |
| sometimes | 9 | 103 | 0.08% | sometime, sometimes |
| future | 6 | 103 | 0.08% | future |
| gone | 4 | 103 | 0.08% | gone |
| headaches | 9 | 103 | 0.08% | headache, headaches |
| something | 9 | 103 | 0.08% | something |
| husband | 7 | 103 | 0.08% | husband, husbands |
| throat | 6 | 103 | 0.08% | throat, throats |
| points | 6 | 102 | 0.08% | point, pointed, points |
| away | 4 | 101 | 0.08% | away |
| readings | 8 | 101 | 0.08% | read, reading, readings |
| consultant | 10 | 101 | 0.08% | consultant, consultants, consultation, consultations, consulted, consulting, consults |
| tell | 4 | 101 | 0.08% | tell, telling, tells |
| actually | 8 | 100 | 0.07% | actual, actually |
| however | 7 | 100 | 0.07% | however |
| bit | 3 | 99 | 0.07% | bit, bits |
| previously | 10 | 99 | 0.07% | previous, previously |
| sent | 4 | 98 | 0.07% | sent |
| referred | 8 | 98 | 0.07% | refer, reference, referred, referring |
| aching | 6 | 98 | 0.07% | ache, aches, aching |
| received | 8 | 98 | 0.07% | receive, received, receiving |
| ideas | 5 | 97 | 0.07% | idea, ideas |
| old | 3 | 97 | 0.07% | old, olds |
| permanently | 11 | 96 | 0.07% | permanantly, permanent, permanently |
| another | 7 | 96 | 0.07% | another |
| fully | 5 | 96 | 0.07% | fully |
| seen | 4 | 96 | 0.07% | seen |
| worst | 5 | 96 | 0.07% | worst |
| stop | 4 | 96 | 0.07% | stop, stopped, stopping, stops |
| sore | 4 | 95 | 0.07% | sore, sorely, soreness, sores |
| specialist | 10 | 95 | 0.07% | specialist, specialists |
| terrifying | 10 | 95 | 0.07% | terrified, terrifiing, terrifying |
| slow | 4 | 95 | 0.07% | slow, slowed, slowing, slows |
| increased | 9 | 94 | 0.07% | increase, increased, increases, increasing, increasingly |
| diagnosing | 10 | 93 | 0.07% | diagnosable, diagnose, diagnosed, diagnoses, diagnosing |
| friends | 7 | 93 | 0.07% | friend, friends |
| steroids | 8 | 93 | 0.07% | steroid, steroids |
| dismissive | 10 | 92 | 0.07% | 'dismissed', dismiss, dismissed, dismisses, dismissing, dismissive, dismissiveness |
| despite | 7 | 92 | 0.07% | despite |
| surgery | 7 | 91 | 0.07% | surgeries, surgery |
| covid19 | 7 | 91 | 0.07% | covid19 |
| next | 4 | 90 | 0.07% | next |
| greatly | 7 | 90 | 0.07% | 'great', great, greatly |
| currently | 9 | 89 | 0.07% | current, currently |
| wonder | 6 | 89 | 0.07% | wonder, wondered, wonderful, wondering |
| best | 4 | 89 | 0.07% | best |
| far | 3 | 89 | 0.07% | far |
| nearly | 6 | 89 | 0.07% | near, nearly |
| publicity | 9 | 89 | 0.07% | public, publication, publications, publicity, publicized |
| please | 6 | 89 | 0.07% | please, pleased |
| pneumonia | 9 | 88 | 0.07% | pneumonia |
| always | 6 | 88 | 0.07% | 'always, always |
| low | 3 | 88 | 0.07% | low, lows |
| smell | 5 | 87 | 0.07% | smell, smells |
| ago | 3 | 87 | 0.07% | ago |
| almost | 6 | 87 | 0.07% | almost |
| lost | 4 | 87 | 0.07% | lost |
| took | 4 | 87 | 0.07% | took |
| usually | 7 | 87 | 0.07% | usual, usually |
| wish | 4 | 86 | 0.06% | wish, wished, wishes, wishing |
| realising | 9 | 85 | 0.06% | realisation, realise, realised, realising |
| attacks | 7 | 85 | 0.06% | attack, attacked, attacks |
| job | 3 | 84 | 0.06% | job, jobs |
| poorly | 6 | 84 | 0.06% | poor, poorly |
| temperature | 11 | 84 | 0.06% | temperature, temperatures |
| often | 5 | 84 | 0.06% | often |
| came | 4 | 84 | 0.06% | 'came, came |
| practice | 8 | 83 | 0.06% | practical, practically, practice, practices |
| period | 6 | 83 | 0.06% | period, periodic, periods |
| hospitalised | 12 | 83 | 0.06% | hospitalisation, hospitalised |
| plans | 5 | 82 | 0.06% | plan, planned, planning, plans |
| totally | 7 | 82 | 0.06% | total, totally |
| loss | 4 | 81 | 0.06% | loss, lossing |
| oxygen | 6 | 81 | 0.06% | oxygen |
| disease | 7 | 80 | 0.06% | disease, diseases |
| guidance | 8 | 80 | 0.06% | guidance, guidances |
| confirm | 7 | 79 | 0.06% | confirm, confirmation, confirmed |
| lucky | 5 | 79 | 0.06% | lucky |
| investigations | 14 | 79 | 0.06% | investigate, investigated, investigating, investigation, investigations |
| developing | 10 | 79 | 0.06% | develop, developed, developing, developments, develops |
| shows | 5 | 78 | 0.06% | show, showed, showing, shows |
| online | 6 | 77 | 0.06% | online |
| muscles | 7 | 77 | 0.06% | 'muscle', muscle, muscles |
| course | 6 | 77 | 0.06% | course, courses |
| 'fine' | 6 | 76 | 0.06% | 'fine', fine |
| acknowledgement | 15 | 76 | 0.06% | acknowledge, acknowledged, acknowledgement, acknowledges, acknowledging, acknowledgment, acknowledgments |
| setting | 7 | 76 | 0.06% | set, sets, setting, settings |
| diagnosis | 9 | 76 | 0.06% | diagnosis |
| real | 4 | 76 | 0.06% | real |
| discharge | 9 | 75 | 0.06% | discharge, discharged |
| admitted | 8 | 74 | 0.06% | admit, admits, admitted, admitting |
| healthcare | 10 | 74 | 0.06% | healthcare, healthcarer |
| progress | 8 | 73 | 0.05% | progress, progressed, progresses, progressing, progression, progressive, progressively |
| stressful | 9 | 73 | 0.05% | stress, stressed, stressful, stressing |
| abandoned | 9 | 73 | 0.05% | abandone, abandoned, abandonement, abandonment |
| info | 4 | 72 | 0.05% | info |
| wasnâ | 5 | 72 | 0.05% | wasnâ |
| weakness | 8 | 72 | 0.05% | weak, weakness, weaknesses |
| catch | 5 | 72 | 0.05% | catch, catches, catching |
| prescribing | 11 | 71 | 0.05% | prescribe, prescribed, prescribes, prescribing |
| xray | 4 | 71 | 0.05% | xray, xrays |
| less | 4 | 71 | 0.05% | less |
| slightly | 8 | 70 | 0.05% | slight, slightly |
| everything | 10 | 70 | 0.05% | everything |
| media | 5 | 70 | 0.05% | media |
| similar | 7 | 69 | 0.05% | similar, similarly |
| else | 4 | 69 | 0.05% | else |
| immunity | 8 | 69 | 0.05% | immune, immunity |
| interested | 10 | 69 | 0.05% | interest, interested, interesting |
| particularly | 12 | 69 | 0.05% | particular, particularly |
| house | 5 | 69 | 0.05% | house, houses |
| places | 6 | 69 | 0.05% | place, placed, places |
| telephone | 9 | 68 | 0.05% | telephone, telephoned |
| limiting | 8 | 68 | 0.05% | limit, limitations, limited, limiting, limits |
| sharing | 7 | 68 | 0.05% | share, shared, sharing |
| system | 6 | 68 | 0.05% | system, systemically, systems |
| taste | 5 | 68 | 0.05% | taste, tastes |
| advised | 7 | 68 | 0.05% | advise, advised, advising |
| visit | 5 | 68 | 0.05% | visit, visited, visiting, visits |
| everyone | 8 | 67 | 0.05% | everyone |
| rather | 6 | 67 | 0.05% | rather |
| three | 5 | 67 | 0.05% | three |
| twice | 5 | 67 | 0.05% | twice |
| whole | 5 | 67 | 0.05% | whole |
| explain | 7 | 67 | 0.05% | explain, explained, explaining |
| provide | 7 | 66 | 0.05% | provide, provided, provider, providers, provides, providing |
| assessment | 10 | 66 | 0.05% | assess, assessed, assessing, assessment, assessments |
| exertion | 8 | 65 | 0.05% | exert, exerted, exerting, exertion, exertional, exertions |
| monitor | 7 | 65 | 0.05% | monitor, monitored, monitoring |
| brain | 5 | 64 | 0.05% | brain, brain' |
| eating | 6 | 64 | 0.05% | eat, eating, eats |
| rate | 4 | 64 | 0.05% | rate, rated |
| questions | 9 | 64 | 0.05% | question, questioned, questioning, questions |
| terrible | 8 | 64 | 0.05% | terrible, terribly |
| physio | 6 | 63 | 0.05% | physio, physios |
| pressure | 8 | 63 | 0.05% | pressure, pressured, pressures |
| became | 6 | 63 | 0.05% | became |
| children | 8 | 63 | 0.05% | children |
| parts | 5 | 63 | 0.05% | part, parting, partly, parts |
| moment | 6 | 63 | 0.05% | moment, moments |
| turn | 4 | 63 | 0.05% | turn, turned, turning, turns |
| suggested | 9 | 62 | 0.05% | suggest, suggested, suggesting, suggestion, suggestions, suggestive, suggests |
| knowledgeable | 13 | 62 | 0.05% | knowledgable, knowledge, knowledgeable |
| already | 7 | 62 | 0.05% | already |
| flu | 3 | 62 | 0.05% | 'flu, flu, flu' |
| website | 7 | 61 | 0.05% | website, websites |
| whilst | 6 | 61 | 0.05% | whilst |
| unknown | 7 | 61 | 0.05% | unknown, unknowns |
| couple | 6 | 61 | 0.05% | couple, coupled |
| stairs | 6 | 61 | 0.05% | stair, stairs |
| heading | 7 | 61 | 0.05% | head, head', headed, heading |
| joint | 5 | 60 | 0.04% | joint, joints |
| quickly | 7 | 60 | 0.04% | quick, quickly |
| reduced | 7 | 60 | 0.04% | reduce, reduced, reducing |
| ambulance | 9 | 60 | 0.04% | ambulance, ambulances |
| anyone | 6 | 60 | 0.04% | anyone, anyones |
| known | 5 | 60 | 0.04% | known |
| staff | 5 | 59 | 0.04% | staff, staffs |
| rehabilitation | 14 | 59 | 0.04% | rehabilitate, rehabilitated, rehabilitating, rehabilitation |
| dont | 4 | 59 | 0.04% | dont |
| slowly | 6 | 59 | 0.04% | slowly |
| recognised | 10 | 59 | 0.04% | recognise, recognised, recognises, recognising, recognision |
| definitely | 10 | 59 | 0.04% | definite, definitely, definition, definitive, definitly |
| including | 9 | 58 | 0.04% | include, included, includes, including |
| april | 5 | 58 | 0.04% | april |
| least | 5 | 58 | 0.04% | least |
| appear | 6 | 58 | 0.04% | appear, appearance, appeared, appearing, appears |
| pushing | 7 | 58 | 0.04% | 'push, push, pushed, pushing |
| function | 8 | 58 | 0.04% | function, functional, functioning, functions |
| main | 4 | 58 | 0.04% | main, mainly |
| havenâ | 6 | 57 | 0.04% | havenâ |
| emotions | 8 | 57 | 0.04% | emotion, emotional, emotionally, emotions |
| burning | 7 | 57 | 0.04% | burn, burning, burns |
| waves | 5 | 56 | 0.04% | wave, waves |
| regards | 7 | 55 | 0.04% | regard, regarded, regarding, regards |
| easily | 6 | 55 | 0.04% | easily |
| sense | 5 | 55 | 0.04% | sense |
| locally | 7 | 54 | 0.04% | local, locality, locally |
| cold | 4 | 54 | 0.04% | cold, colds |
| ups | 3 | 54 | 0.04% | up', upped, ups |
| tiredness | 9 | 53 | 0.04% | tiredness |
| kind | 4 | 53 | 0.04% | kind, kindly, kindness |
| means | 5 | 53 | 0.04% | mean, meaning, means |
| discuss | 7 | 53 | 0.04% | discuss, discussed, discussing, discussion, discussions |
| peak | 4 | 53 | 0.04% | peak, peaked, peaking, peaks |
| wake | 4 | 53 | 0.04% | wake, wakes, waking |
| beginning | 9 | 52 | 0.04% | begin, begining, beginning, beginnings |
| focus | 5 | 52 | 0.04% | focus, focused, focuses, focusing |
| suddenly | 8 | 52 | 0.04% | sudden, suddenly |
| swab | 4 | 52 | 0.04% | swab, swabbed, swabs |
| team | 4 | 52 | 0.04% | team, teamed, teams |
| grateful | 8 | 52 | 0.04% | grateful |
| huge | 4 | 51 | 0.04% | huge, hugely |
| lingering | 9 | 51 | 0.04% | linger, lingered, lingering, lingers |
| control | 7 | 51 | 0.04% | control, controlled, controlling |
| intensive | 9 | 51 | 0.04% | intense, intensely, intensity, intensive |
| second | 6 | 51 | 0.04% | second, secondly |
| moving | 6 | 51 | 0.04% | move, moved, moves, moving |
| present | 7 | 51 | 0.04% | present, presentation, presentations, presented, presenters, presenting, presently |
| let | 3 | 51 | 0.04% | let, letting |
| finally | 7 | 50 | 0.04% | 'finally, final, finally |
| nightmare | 9 | 50 | 0.04% | nightmare, nightmares |
| pace | 4 | 50 | 0.04% | pace, pacing |
| sats | 4 | 50 | 0.04% | sat, sats |
| wrong | 5 | 50 | 0.04% | wrong, wrongly |
| horrible | 8 | 50 | 0.04% | horrible |
| maybe | 5 | 50 | 0.04% | maybe |
| recognition | 11 | 50 | 0.04% | recognition |
| unsure | 6 | 50 | 0.04% | unsure |
| persistent | 10 | 50 | 0.04% | persist, persistance, persisted, persistent, persisting, persists |
| probably | 8 | 50 | 0.04% | probability, probable, probably |
| side | 4 | 50 | 0.04% | side, sided, sides |
| traumatic | 9 | 50 | 0.04% | traumatic, traumatized, traumatizing |
| chronic | 7 | 49 | 0.04% | chronic, chronically |
| caught | 6 | 49 | 0.04% | caught |
| couldnâ | 7 | 49 | 0.04% | couldnâ |
| fog | 3 | 49 | 0.04% | fog |
| half | 4 | 49 | 0.04% | half |
| minutes | 7 | 49 | 0.04% | minute, minutes |
| appreciate | 10 | 49 | 0.04% | appreciate, appreciated, appreciating, appreciation, appreciative |
| dizziness | 9 | 49 | 0.04% | dizziness, dizzy |
| facebook | 8 | 48 | 0.04% | facebook |
| passing | 7 | 48 | 0.04% | 'pass, pass, passed, passes, passing |
| busy | 4 | 48 | 0.04% | business, businesses, busy |
| incredibly | 10 | 48 | 0.04% | incredible, incredibly |
| learning | 8 | 47 | 0.04% | learn, learned, learning |
| cycling | 7 | 47 | 0.04% | cycle, cycles, cycling |
| horrendous | 10 | 47 | 0.04% | horrendous |
| lockdown | 8 | 47 | 0.04% | lockdown |
| hold | 4 | 47 | 0.04% | hold, holding, holds |
| list | 4 | 46 | 0.03% | list, listed, listing |
| shopping | 8 | 46 | 0.03% | shop, shopping, shops |
| community | 9 | 46 | 0.03% | communities, community |
| unless | 6 | 46 | 0.03% | 'unless, unless |
| cant | 4 | 46 | 0.03% | cant |
| nobody | 6 | 46 | 0.03% | nobody |
| none | 4 | 46 | 0.03% | none |
| required | 8 | 46 | 0.03% | require, required, requirements, requires, requiring |
| risk | 4 | 46 | 0.03% | risk, risked, risking, risks |
| goes | 4 | 45 | 0.03% | goes |
| instead | 7 | 45 | 0.03% | instead |
| rehab | 5 | 45 | 0.03% | rehab |
| sob | 3 | 45 | 0.03% | sob |
| soon | 4 | 45 | 0.03% | soon |
| hit | 3 | 45 | 0.03% | hit, hits, hitting |
| socially | 8 | 45 | 0.03% | social, socially |
| upsetting | 9 | 45 | 0.03% | upset, upsets, upsetting |
| contracting | 11 | 45 | 0.03% | contract, contracted, contracting |
| suspected | 9 | 45 | 0.03% | suspect, suspected, suspectedly, suspecting, suspects |
| anti | 4 | 44 | 0.03% | anti |
| apart | 5 | 44 | 0.03% | apart |
| doesnâ | 6 | 44 | 0.03% | doesnâ |
| either | 6 | 44 | 0.03% | either |
| knew | 4 | 44 | 0.03% | knew |
| paramedics | 10 | 44 | 0.03% | paramedic, paramedics |
| ignoring | 8 | 44 | 0.03% | ignorance, ignorant, ignore, ignored, ignores, ignoring |
| carrying | 8 | 43 | 0.03% | carried, carries, carry, carrying |
| phased | 6 | 43 | 0.03% | phase, phased, phases, phasing |
| number | 6 | 43 | 0.03% | number, numbers |
| sit | 3 | 43 | 0.03% | sit, sitting |
| gave | 4 | 43 | 0.03% | gave |
| ability | 7 | 42 | 0.03% | abilities, ability |
| fighting | 8 | 42 | 0.03% | 'fight, fight, fighting |
| mind | 4 | 42 | 0.03% | mind, mindful, mindfully, minds |
| reason | 6 | 42 | 0.03% | reason, reasonable, reasonably, reasoned, reasons |
| 100 | 3 | 42 | 0.03% | 100 |
| legs | 4 | 42 | 0.03% | leg, legs |
| plus | 4 | 42 | 0.03% | plus |
| pretty | 6 | 42 | 0.03% | pretty |
| saying | 6 | 42 | 0.03% | saying |
| shielding | 9 | 42 | 0.03% | shield, shielded, shielding |
| hubs | 4 | 42 | 0.03% | hub, hubs |
| tasks | 5 | 42 | 0.03% | task, tasks |
| situation | 9 | 41 | 0.03% | situation, situations |
| weight | 6 | 41 | 0.03% | weight, weights |
| desperate | 9 | 41 | 0.03% | desperate, desperately |
| financial | 9 | 41 | 0.03% | financial, financially |
| morning | 7 | 41 | 0.03% | morning, mornings |
| uncertain | 9 | 41 | 0.03% | uncertain |
| hands | 5 | 41 | 0.03% | hand, handed, handfuls, hands |
| examination | 11 | 41 | 0.03% | examination, examinations, examine, examined |
| recently | 8 | 41 | 0.03% | recent, recently |
| triggered | 9 | 41 | 0.03% | trigger, triggered, triggering, triggers |
| communication | 13 | 41 | 0.03% | communicate, communicated, communication, communications |
| flow | 4 | 41 | 0.03% | flow, flowed, flowing, flows |
| referral | 8 | 41 | 0.03% | referral, referrals |
| remains | 7 | 41 | 0.03% | remain, remained, remaining, remains |
| kept | 4 | 40 | 0.03% | kept |
| pre | 3 | 40 | 0.03% | pre |
| prior | 5 | 40 | 0.03% | prior |
| throughout | 10 | 40 | 0.03% | throughout |
| young | 5 | 40 | 0.03% | young |
| late | 4 | 40 | 0.03% | late, lately |
| surviving | 9 | 40 | 0.03% | 'surviving', survive, survived, surviving |
| response | 8 | 40 | 0.03% | response, responses, responsibility, responsible, responsive |
| absolutely | 10 | 40 | 0.03% | absolute, absolutely |
| moderate | 8 | 40 | 0.03% | moderate, moderately |
| challenging | 11 | 39 | 0.03% | challenge, challenges, challenging |
| daughter | 8 | 39 | 0.03% | daughter, daughters |
| ages | 4 | 39 | 0.03% | age, aged, ageing, ages |
| significantly | 13 | 39 | 0.03% | significance, significant, significantly |
| love | 4 | 39 | 0.03% | love, loved, lovely |
| rang | 4 | 39 | 0.03% | rang, range, ranged, ranging |
| signs | 5 | 39 | 0.03% | sign, signed, signing, signs |
| mask | 4 | 39 | 0.03% | mask, masked, masking, masks |
| strangely | 9 | 39 | 0.03% | strange, strangely |
| certain | 7 | 39 | 0.03% | certain, certainly |
| distressing | 11 | 39 | 0.03% | distress, distressed, distressing |
| missed | 6 | 39 | 0.03% | miss, missed, missing |
| profession | 10 | 39 | 0.03% | profession, professions |
| concentrate | 11 | 39 | 0.03% | concentrate, concentrated, concentrating, concentration |
| surprising | 10 | 39 | 0.03% | surprise, surprised, surprising, surprisingly |
| emerging | 8 | 39 | 0.03% | emerge, emerged, emergencies, emergency, emerges, emerging |
| line | 4 | 39 | 0.03% | line, lined, lines |
| small | 5 | 39 | 0.03% | small |
| wife | 4 | 39 | 0.03% | wife |
| yes | 3 | 39 | 0.03% | yes |
| admission | 9 | 39 | 0.03% | admission, admissions |
| mood | 4 | 39 | 0.03% | mood, moods |
| forward | 7 | 38 | 0.03% | forward, forwarded, forwards |
| facts | 5 | 38 | 0.03% | fact, facts |
| seek | 4 | 38 | 0.03% | seek, seeking |
| worsening | 9 | 38 | 0.03% | worsen, worsened, worsening, worsens |
| close | 5 | 38 | 0.03% | close, closed, closely, closing |
| capacity | 8 | 38 | 0.03% | capacity |
| hear | 4 | 38 | 0.03% | hear, hearing |
| prolonged | 9 | 38 | 0.03% | prolonged |
| vulnerable | 10 | 38 | 0.03% | vulnerability, vulnerable |
| important | 9 | 38 | 0.03% | import, importance, important, importantly |
| existent | 8 | 37 | 0.03% | exist, existant, existed, existence, existent, existing, exists |
| 2020 | 4 | 37 | 0.03% | 2020 |
| nice | 4 | 37 | 0.03% | nice |
| pay | 3 | 37 | 0.03% | pay, paying |
| pulmonary | 9 | 37 | 0.03% | pulmonary |
| relieved | 8 | 37 | 0.03% | relieve, relieved, reliever |
| secondary | 9 | 37 | 0.03% | secondary |
| wheezing | 8 | 37 | 0.03% | wheeze, wheezing |
| plays | 5 | 37 | 0.03% | play, played, playful, playing, plays |
| accept | 6 | 37 | 0.03% | accept, acceptable, acceptance, accepted, accepting |
| review | 6 | 37 | 0.03% | review, reviewed, reviewing, reviews |
| considering | 11 | 37 | 0.03% | consider, considered, considering |
| lightly | 7 | 37 | 0.03% | light, lightly |
| sadness | 7 | 37 | 0.03% | sad, sadly, sadness |
| must | 4 | 36 | 0.03% | must |
| past | 4 | 36 | 0.03% | past |
| massive | 7 | 36 | 0.03% | massive, massively |
| occasions | 9 | 36 | 0.03% | occasion, occasions |
| sensation | 9 | 36 | 0.03% | sensation, sensations |
| type | 4 | 36 | 0.03% | type, typed, types |
| link | 4 | 36 | 0.03% | link, linked, linking, links |
| private | 7 | 36 | 0.03% | private, privately |
| air | 3 | 36 | 0.03% | air, aired |
| losing | 6 | 35 | 0.03% | lose, losing |
| noticing | 8 | 35 | 0.03% | noticable, notice, noticeable, noticed, noticing |
| garden | 6 | 35 | 0.03% | garden, gardened, gardener, gardening, gardens |
| stage | 5 | 35 | 0.03% | stage, stages |
| barely | 6 | 35 | 0.03% | bare, barely |
| survey | 6 | 35 | 0.03% | survey, surveys |
| tail | 4 | 35 | 0.03% | tail, tail', tailed |
| deep | 4 | 35 | 0.03% | deep |
| spoke | 5 | 35 | 0.03% | spoke |
| unsupported | 11 | 35 | 0.03% | unsupported |
| within | 6 | 35 | 0.03% | within |
| words | 5 | 34 | 0.03% | word, wording, words |
| repeats | 7 | 34 | 0.03% | repeat, repeated, repeatedly, repeating, repeats |
| thereâ | 6 | 34 | 0.03% | thereâ |
| uncertainty | 11 | 34 | 0.03% | uncertainties, uncertainty |
| gradually | 9 | 33 | 0.02% | gradual, gradually |
| angry | 5 | 33 | 0.02% | angry |
| drs | 3 | 33 | 0.02% | drs |
| eventually | 10 | 33 | 0.02% | eventually |
| everyday | 8 | 33 | 0.02% | everyday |
| four | 4 | 33 | 0.02% | four |
| inflammation | 12 | 33 | 0.02% | inflammation |
| ring | 4 | 33 | 0.02% | ring, ringing |
| wearing | 7 | 33 | 0.02% | wear, wearing, wears |
| ease | 4 | 33 | 0.02% | ease, eased, eases, easing |
| except | 6 | 33 | 0.02% | except, exception |
| infectious | 10 | 33 | 0.02% | infectious, infectiousness |
| sort | 4 | 33 | 0.02% | sort, sorted, sorting, sorts |
| allowed | 7 | 33 | 0.02% | allow, allowed, allowing, allows |
| happy | 5 | 33 | 0.02% | happiness, happy |
| strength | 8 | 33 | 0.02% | strength, strengths |
| distancing | 10 | 33 | 0.02% | distance, distanced, distances, distancing |
| form | 4 | 33 | 0.02% | form, formed, forms |
| heavy | 5 | 32 | 0.02% | heaviness, heavy |
| assuming | 8 | 32 | 0.02% | assume, assumed, assumes, assuming |
| shocking | 8 | 32 | 0.02% | shock, shocked, shocking, shockingly |
| basically | 9 | 32 | 0.02% | basic, basically |
| drop | 4 | 32 | 0.02% | drop, dropped, dropping, drops |
| note | 4 | 32 | 0.02% | note, notes |
| obviously | 9 | 32 | 0.02% | obvious, obviously |
| strongly | 8 | 32 | 0.02% | strong, strongly |
| afterwards | 10 | 32 | 0.02% | afterward, afterwards |
| earlier | 7 | 32 | 0.02% | 'earlier', earlier |
| example | 7 | 32 | 0.02% | example, examples |
| haul | 4 | 32 | 0.02% | haul, haul' |
| build | 5 | 32 | 0.02% | build, building, buildings, builds |
| complications | 13 | 32 | 0.02% | complicated, complicating, complication, complications |
| disappointed | 12 | 32 | 0.02% | disappointed, disappointing, disappointment |
| bother | 6 | 32 | 0.02% | bother, bothered, bothering, bothers |
| drink | 5 | 32 | 0.02% | drink, drinking, drinks |
| date | 4 | 32 | 0.02% | date, dates |
| non | 3 | 32 | 0.02% | non |
| via | 3 | 32 | 0.02% | via |
| food | 4 | 32 | 0.02% | food, foods |
| safe | 4 | 32 | 0.02% | safe, safely |
| miles | 5 | 31 | 0.02% | mile, miles |
| worker | 6 | 31 | 0.02% | worker, workers |
| resources | 9 | 31 | 0.02% | resource, resources |
| asthmatic | 9 | 31 | 0.02% | asthmatic, asthmatics |
| big | 3 | 31 | 0.02% | big |
| dry | 3 | 31 | 0.02% | dry |
| forgotten | 9 | 31 | 0.02% | forgotten |
| partner | 7 | 31 | 0.02% | partner |
| rollercoaster | 13 | 31 | 0.02% | rollercoaster |
| today | 5 | 31 | 0.02% | today |
| attended | 8 | 31 | 0.02% | attend, attended, attending |
| stating | 7 | 31 | 0.02% | state, stated, states, stating |
| avoid | 5 | 31 | 0.02% | avoid, avoidable, avoided, avoiding |
| lying | 5 | 31 | 0.02% | lie, lied, lies, lying |
| wards | 5 | 31 | 0.02% | ward, wards |
| glad | 4 | 31 | 0.02% | glad, gladly |
| mention | 7 | 31 | 0.02% | mention, mentioned, mentioning |
| large | 5 | 30 | 0.02% | large, largely |
| flares | 6 | 30 | 0.02% | 'flare, flare, flared, flares |
| prevent | 7 | 30 | 0.02% | prevent, preventative, prevented, preventer, preventing, prevention, prevents |
| confidence | 10 | 30 | 0.02% | confidence, confident, confidently |
| length | 6 | 30 | 0.02% | length, lengths |
| process | 7 | 30 | 0.02% | process, processed |
| describe | 8 | 30 | 0.02% | describe, described, describes, describing |
| specific | 8 | 30 | 0.02% | specific, specifically |
| heard | 5 | 30 | 0.02% | heard |
| hot | 3 | 30 | 0.02% | hot |
| palpitations | 12 | 30 | 0.02% | palpitations |
| panic | 5 | 30 | 0.02% | panic |
| sons | 4 | 30 | 0.02% | son, sons |
| steps | 5 | 29 | 0.02% | step, steps |
| along | 5 | 29 | 0.02% | along |
| area | 4 | 29 | 0.02% | area, areas |
| coronavirus | 11 | 29 | 0.02% | coronavirus |
| fed | 3 | 29 | 0.02% | fed |
| impossible | 10 | 29 | 0.02% | impossible |
| june | 4 | 29 | 0.02% | june |
| spoken | 6 | 29 | 0.02% | spoken |
| therefore | 9 | 29 | 0.02% | therefore |
| understood | 10 | 29 | 0.02% | understood |
| haulers | 7 | 29 | 0.02% | hauler, haulers, haulers' |
| watching | 8 | 29 | 0.02% | 'watching, watch, watched, watchful, watching |
| openly | 6 | 29 | 0.02% | open, opened, opening, openly, openness |
| refusing | 8 | 29 | 0.02% | refuse, refused, refuses, refusing |
| enjoy | 5 | 29 | 0.02% | enjoy, enjoyable, enjoyed, enjoying, enjoyment |
| eye | 3 | 29 | 0.02% | eye, eyes |
| leads | 5 | 29 | 0.02% | lead, leading, leads |
| clots | 5 | 28 | 0.02% | clot, clots, clotting |
| dog | 3 | 28 | 0.02% | dog, dogs, dogs' |
| colleagues | 10 | 28 | 0.02% | colleague, colleagues |
| restricted | 10 | 28 | 0.02% | restricted, restricting, restrictions, restrictive, restricts |
| ive | 3 | 28 | 0.02% | ive, ives |
| amazingly | 9 | 28 | 0.02% | amazes, amazing, amazingly |
| acute | 5 | 28 | 0.02% | acute, acutely |
| school | 6 | 28 | 0.02% | school, schools |
| joining | 7 | 28 | 0.02% | 'joined, join, joined, joining |
| major | 5 | 28 | 0.02% | major, majority, majors |
| alive | 5 | 28 | 0.02% | alive |
| mostly | 6 | 28 | 0.02% | mostly |
| saw | 3 | 28 | 0.02% | saw |
| simple | 6 | 28 | 0.02% | simple |
| voice | 5 | 28 | 0.02% | voice |
| ecg | 3 | 28 | 0.02% | ecg, ecgs |
| compared | 8 | 27 | 0.02% | comparable, comparatively, compare, compared, comparing |
| appetite | 8 | 27 | 0.02% | appetite |
| horrific | 8 | 27 | 0.02% | horrific |
| paracetamol | 11 | 27 | 0.02% | paracetamol |
| syndrome | 8 | 27 | 0.02% | syndrome |
| raised | 6 | 27 | 0.02% | raise, raised, raises, raising |
| disabled | 8 | 27 | 0.02% | disability, disabled, disabling |
| falling | 7 | 27 | 0.02% | fall, falling, falls |
| pandemic | 8 | 27 | 0.02% | pandemic, pandemics |
| education | 9 | 26 | 0.02% | educate, educated, educating, education |
| appropriate | 11 | 26 | 0.02% | appropriate, appropriately |
| studying | 8 | 26 | 0.02% | studied, studies, study, studying |
| draining | 8 | 26 | 0.02% | drained, draining |
| varied | 6 | 26 | 0.02% | varied, varies, vary, varying |
| honestly | 8 | 26 | 0.02% | honest, honestly |
| parents | 7 | 26 | 0.02% | parent, parents |
| smoke | 5 | 26 | 0.02% | smoke, smoked, smoking |
| standing | 8 | 26 | 0.02% | stand, standing |
| hurts | 5 | 26 | 0.02% | hurt, hurtful, hurting, hurts |
| spend | 5 | 26 | 0.02% | spend, spending, spends |
| battle | 6 | 26 | 0.02% | battle, battled, battles, battling |
| unexpectedly | 12 | 26 | 0.02% | unexpectably, unexpected, unexpectedly |
| february | 8 | 26 | 0.02% | february |
| guilty | 6 | 26 | 0.02% | guilty |
| isnâ | 4 | 26 | 0.02% | isnâ |
| multiple | 8 | 26 | 0.02% | multiple |
| mum | 3 | 26 | 0.02% | mum |
| upper | 5 | 26 | 0.02% | upper |
| wonâ | 4 | 26 | 0.02% | wonâ |
| yoga | 4 | 26 | 0.02% | yoga |
| amount | 6 | 25 | 0.02% | amount, amounts |
| imagining | 9 | 25 | 0.02% | imagination, imagine, imagining |
| memory | 6 | 25 | 0.02% | memories, memory |
| neurology | 9 | 25 | 0.02% | neurological, neurology |
| additional | 10 | 25 | 0.02% | addition, additional, additionally |
| dose | 4 | 25 | 0.02% | dose, doses |
| single | 6 | 25 | 0.02% | single, singled |
| centre | 6 | 25 | 0.02% | centre, centred, centres |
| request | 7 | 25 | 0.02% | request, requested, requesting, requests |
| forgetful | 9 | 25 | 0.02% | forget, forgetful, forgetfulness, forgetting |
| basis | 5 | 25 | 0.02% | basis |
| blue | 4 | 25 | 0.02% | blue |
| cv19 | 4 | 25 | 0.02% | cv19 |
| discomfort | 10 | 25 | 0.02% | discomfort |
| episodes | 8 | 25 | 0.02% | episode, episodes |
| frequent | 8 | 25 | 0.02% | frequent, frequently |
| guidelines | 10 | 25 | 0.02% | guidelines |
| icu | 3 | 25 | 0.02% | icu |
| journey | 7 | 25 | 0.02% | journey |
| nose | 4 | 25 | 0.02% | nose |
| potential | 9 | 25 | 0.02% | potential, potentially |
| annoying | 8 | 25 | 0.02% | annoyance, annoyed, annoying |
| directed | 8 | 25 | 0.02% | direct, directed, directing, direction, directly |
| round | 5 | 25 | 0.02% | round, rounds |
| shower | 6 | 25 | 0.02% | shower, showered, showering, showers |
| conversation | 12 | 25 | 0.02% | conversation, conversations |
| members | 7 | 25 | 0.02% | member, members |
| dreadful | 8 | 25 | 0.02% | dread, dreadful, dreadfully, dreading |
| copd | 4 | 24 | 0.02% | copd |
| easy | 4 | 24 | 0.02% | easy |
| january | 7 | 24 | 0.02% | january |
| nowhere | 7 | 24 | 0.02% | nowhere |
| stomach | 7 | 24 | 0.02% | stomach |
| details | 7 | 24 | 0.02% | detail, detailed, details |
| therapy | 7 | 24 | 0.02% | therapies, therapy |
| individual | 10 | 24 | 0.02% | individual, individually, individuals |
| temp | 4 | 24 | 0.02% | temp, temps |
| ride | 4 | 24 | 0.02% | ride, rides, riding |
| aid | 3 | 24 | 0.02% | aid, aide, aided, aiding, aids |
| books | 5 | 24 | 0.02% | book, booked, booking, books |
| vitamins | 8 | 24 | 0.02% | vitamin, vitamins |
| ensure | 6 | 23 | 0.02% | ensure, ensuring |
| news | 4 | 23 | 0.02% | news, news' |
| pulse | 5 | 23 | 0.02% | pulse, pulses |
| doubt | 5 | 23 | 0.02% | doubt, doubted, doubtful, doubting, doubts |
| training | 8 | 23 | 0.02% | train, trained, training, trains |
| updates | 7 | 23 | 0.02% | update, updated, updates |
| assistance | 10 | 23 | 0.02% | assist, assistance, assistant, assistants |
| sound | 5 | 23 | 0.02% | sound, sounded, sounding, soundly, sounds |
| 999 | 3 | 23 | 0.02% | 999 |
| afraid | 6 | 23 | 0.02% | afraid |
| fast | 4 | 23 | 0.02% | fast |
| gpâ | 3 | 23 | 0.02% | gpâ |
| relief | 6 | 23 | 0.02% | relief |
| simply | 6 | 23 | 0.02% | simply |
| spent | 5 | 23 | 0.02% | spent |
| tachycardia | 11 | 23 | 0.02% | tachycardia |
| weird | 5 | 23 | 0.02% | weird |
| world | 5 | 23 | 0.02% | world |
| email | 5 | 23 | 0.02% | email, emailed, emailing, emails |
| fill | 4 | 23 | 0.02% | fill, filled, filling, fills |
| rule | 4 | 23 | 0.02% | rule, ruled, rules, ruling |
| sweats | 6 | 23 | 0.02% | sweat, sweated, sweating, sweats |
| effort | 6 | 23 | 0.02% | effort, efforts |
| practitioner | 12 | 23 | 0.02% | practitioner, practitioners |
| send | 4 | 23 | 0.02% | send, sending, sends |
| aftercare | 9 | 22 | 0.02% | aftercare |
| hell | 4 | 22 | 0.02% | hell |
| onset | 5 | 22 | 0.02% | onset |
| primary | 7 | 22 | 0.02% | primary |
| six | 3 | 22 | 0.02% | six |
| somewhat | 8 | 22 | 0.02% | somewhat |
| sympathetic | 11 | 22 | 0.02% | sympathetic |
| underlying | 10 | 22 | 0.02% | underlying |
| various | 7 | 22 | 0.02% | various |
| exactly | 7 | 22 | 0.02% | exact, exacted, exactly |
| cry | 3 | 22 | 0.02% | cried, cry, crying |
| intermittent | 12 | 22 | 0.02% | intermittent, intermittently |
| fairly | 6 | 22 | 0.02% | fair, fairly |
| programme | 9 | 22 | 0.02% | programme, programmes |
| trip | 4 | 22 | 0.02% | trip, trips |
| based | 5 | 22 | 0.02% | base, based, bases |
| exacerbation | 12 | 22 | 0.02% | exacerbate, exacerbated, exacerbates, exacerbation, exacerbations |
| shoulder | 8 | 22 | 0.02% | shoulder, shoulders |
| death | 5 | 21 | 0.02% | death, deaths |
| widely | 6 | 21 | 0.02% | wide, widely |
| false | 5 | 21 | 0.02% | false, falsely |
| fell | 4 | 21 | 0.02% | fell, felling |
| recurring | 9 | 21 | 0.02% | recur, recurred, recurring |
| organised | 9 | 21 | 0.02% | organisation, organisations, organise, organised, organising |
| washing | 7 | 21 | 0.02% | wash, washed, washes, washing |
| benefits | 8 | 21 | 0.02% | benefit, benefited, benefits |
| brings | 6 | 21 | 0.02% | bring, bringing, brings |
| attention | 9 | 21 | 0.02% | attention |
| cancer | 6 | 21 | 0.02% | cancer |
| empathy | 7 | 21 | 0.02% | empathy |
| excellent | 9 | 21 | 0.02% | excellent |
| meant | 5 | 21 | 0.02% | meant |
| mouth | 5 | 21 | 0.02% | mouth |
| psychological | 13 | 21 | 0.02% | psychological, psychologically |
| room | 4 | 21 | 0.02% | room |
| site | 4 | 21 | 0.02% | site, sites |
| thatâ | 5 | 21 | 0.02% | thatâ |
| together | 8 | 21 | 0.02% | together |
| tough | 5 | 21 | 0.02% | tough |
| useless | 7 | 21 | 0.02% | useless |
| teacher | 7 | 21 | 0.02% | teacher, teachers |
| rash | 4 | 21 | 0.02% | rash, rashes |
| racing | 6 | 21 | 0.02% | 'racing', races, racing |
| hill | 4 | 21 | 0.02% | hill, hills |
| apparent | 8 | 20 | 0.01% | apparent, apparently |
| child | 5 | 20 | 0.01% | child |
| feet | 4 | 20 | 0.01% | feet |
| kidney | 6 | 20 | 0.01% | kidney, kidneys |
| meds | 4 | 20 | 0.01% | meds |
| mid | 3 | 20 | 0.01% | mid |
| mucus | 5 | 20 | 0.01% | mucus |
| necessary | 9 | 20 | 0.01% | necessary |
| nervous | 7 | 20 | 0.01% | nervous |
| outside | 7 | 20 | 0.01% | outside |
| pleurisy | 8 | 20 | 0.01% | pleurisy |
| ppe | 3 | 20 | 0.01% | ppe |
| thousands | 9 | 20 | 0.01% | thousands |
| youâ | 4 | 20 | 0.01% | youâ |
| zero | 4 | 20 | 0.01% | zero |
| fobbing | 7 | 20 | 0.01% | 'fobbed, fob, fobbed, fobbing |
| gasping | 7 | 20 | 0.01% | gasp, gasping |
| scarring | 8 | 20 | 0.01% | 'scarring', scar, scarred, scarring |
| meet | 4 | 20 | 0.01% | meet, meeting, meetings |
| deteriorating | 13 | 20 | 0.01% | deteriorate, deteriorated, deteriorating, deterioration |
| indicate | 8 | 20 | 0.01% | indicate, indicated, indicates, indicating, indication, indications, indicative, indicator |
| resolve | 7 | 20 | 0.01% | resolve, resolved, resolves, resolving |
| reports | 7 | 20 | 0.01% | report, reported, reporting, reports |
| cleaning | 8 | 20 | 0.01% | clean, cleaning |
| gentle | 6 | 20 | 0.01% | gentle, gently |
| touch | 5 | 20 | 0.01% | touch, touched |
| gym | 3 | 20 | 0.01% | gym, gyms |
| uncomfortable | 13 | 20 | 0.01% | uncomfortable, uncomfortably |
| critical | 8 | 20 | 0.01% | critical, critically, criticism, critics |
| order | 5 | 20 | 0.01% | order, ordered, ordering |
| unpredictable | 13 | 20 | 0.01% | unpredictability, unpredictable, unpredictably |
| depends | 7 | 20 | 0.01% | depend, dependance, dependancy, dependant, depended, dependency, dependent, depending, depends |
| organs | 6 | 20 | 0.01% | organ, organs |
| routine | 7 | 20 | 0.01% | routine, routinely, routines |
| preparing | 9 | 20 | 0.01% | preparation, prepare, prepared, preparing |
| knocks | 6 | 20 | 0.01% | knock, knocked, knocking, knocks |
| accurate | 8 | 19 | 0.01% | accurate, accurately |
| common | 6 | 19 | 0.01% | common, commonly |
| opportunity | 11 | 19 | 0.01% | opportunities, opportunity |
| text | 4 | 19 | 0.01% | text, texting |
| ventolin | 8 | 19 | 0.01% | ventolin, ventoline |
| counting | 8 | 19 | 0.01% | count, counted, counting |
| ears | 4 | 19 | 0.01% | ear, ears |
| occurring | 9 | 19 | 0.01% | occur, occuring, occurred, occurring, occurs |
| anymore | 7 | 19 | 0.01% | anymore |
| began | 5 | 19 | 0.01% | began |
| dark | 4 | 19 | 0.01% | dark |
| didnt | 5 | 19 | 0.01% | didnt |
| housework | 9 | 19 | 0.01% | housework |
| marathon | 8 | 19 | 0.01% | marathon, marathons |
| stuck | 5 | 19 | 0.01% | stuck |
| top | 3 | 19 | 0.01% | top |
| matter | 6 | 19 | 0.01% | matter, mattered, matters |
| household | 9 | 19 | 0.01% | household, households |
| pattern | 7 | 19 | 0.01% | pattern, patterns |
| fluid | 5 | 19 | 0.01% | fluid, fluids |
| proactive | 9 | 19 | 0.01% | proactive, proactively |
| heal | 4 | 19 | 0.01% | heal, healed, healing |
| anyway | 6 | 18 | 0.01% | anyway |
| arrange | 7 | 18 | 0.01% | arrange, arranged, arranging |
| beyond | 6 | 18 | 0.01% | beyond |
| coaster | 7 | 18 | 0.01% | coaster |
| fortunate | 9 | 18 | 0.01% | fortunate, fortunately |
| front | 5 | 18 | 0.01% | front |
| guilt | 5 | 18 | 0.01% | guilt |
| london | 6 | 18 | 0.01% | london |
| ribs | 4 | 18 | 0.01% | rib, ribs |
| shift | 5 | 18 | 0.01% | shift, shifts |
| skin | 4 | 18 | 0.01% | skin |
| tablets | 7 | 18 | 0.01% | tablets |
| wasting | 7 | 18 | 0.01% | waste, wasted, wasting |
| wider | 5 | 18 | 0.01% | wider |
| climbing | 8 | 18 | 0.01% | climb, climbed, climbing |
| originally | 10 | 18 | 0.01% | origin, original, originally |
| convinced | 9 | 18 | 0.01% | convince, convinced, convincing |
| recommended | 11 | 18 | 0.01% | recommend, recommendation, recommendations, recommended, recommending |
| fluctuating | 11 | 18 | 0.01% | fluctuate, fluctuated, fluctuates, fluctuating, fluctuations |
| consistently | 12 | 18 | 0.01% | consistant, consisted, consistency, consistent, consistently |
| country | 7 | 18 | 0.01% | countries, country |
| stories | 7 | 18 | 0.01% | stories, story |
| qualify | 7 | 18 | 0.01% | qualified, qualify, qualifying |
| swim | 4 | 18 | 0.01% | swim, swimming, swims |
| pick | 4 | 18 | 0.01% | pick, picked, picking |
| chat | 4 | 18 | 0.01% | chat, chatted |
| hair | 4 | 18 | 0.01% | hair, hairs |
| official | 8 | 18 | 0.01% | 'officially', official, officially |
| ten | 3 | 18 | 0.01% | ten, tens |
| cook | 4 | 18 | 0.01% | cook, cooked, cooking |
| mine | 4 | 18 | 0.01% | mine, mines |
| trouble | 7 | 18 | 0.01% | trouble, troubled, troubles, troubling |
| unfortunately | 13 | 18 | 0.01% | unfortunate, unfortunately |
| cognitively | 11 | 18 | 0.01% | cognition, cognitive, cognitively |
| diet | 4 | 18 | 0.01% | diet, dieting, diets |
| experts | 7 | 18 | 0.01% | expert, experts, experts' |
| hopeless | 8 | 18 | 0.01% | hopeless, hopelessly, hopelessness |
| traumatised | 11 | 18 | 0.01% | traumatised, traumatises, traumatising |
| fingers | 7 | 18 | 0.01% | finger, fingers |
| reluctant | 9 | 18 | 0.01% | reluctance, reluctant |
| urgent | 6 | 18 | 0.01% | urgent, urgently |
| spread | 6 | 17 | 0.01% | spread, spreading |
| video | 5 | 17 | 0.01% | video, videos |
| message | 7 | 17 | 0.01% | message, messages, messaging |
| decided | 7 | 17 | 0.01% | decide, decided, deciding |
| arms | 4 | 17 | 0.01% | arm, arms |
| driving | 7 | 17 | 0.01% | drive, driving |
| options | 7 | 17 | 0.01% | option, options |
| ventilator | 10 | 17 | 0.01% | ventilated, ventilating, ventilation, ventilator, ventilators |
| crash | 5 | 17 | 0.01% | crash, crashed, crashing |
| prove | 5 | 17 | 0.01% | prove, proved, proving |
| arenâ | 5 | 17 | 0.01% | arenâ |
| awaiting | 8 | 17 | 0.01% | awaiting |
| cardiac | 7 | 17 | 0.01% | cardiac |
| easier | 6 | 17 | 0.01% | easier |
| kids | 4 | 17 | 0.01% | kids |
| per | 3 | 17 | 0.01% | per |
| ptsd | 4 | 17 | 0.01% | ptsd |
| receptionist | 12 | 17 | 0.01% | receptionist, receptionists |
| reliable | 8 | 17 | 0.01% | reliable |
| roller | 6 | 17 | 0.01% | roller |
| contagious | 10 | 17 | 0.01% | contagious, contagiousness |
| neck | 4 | 17 | 0.01% | neck, neck' |
| odd | 3 | 17 | 0.01% | odd, oddly, odds |
| address | 7 | 17 | 0.01% | address, addressed, addresses, addressing |
| approach | 8 | 17 | 0.01% | approach, approaching |

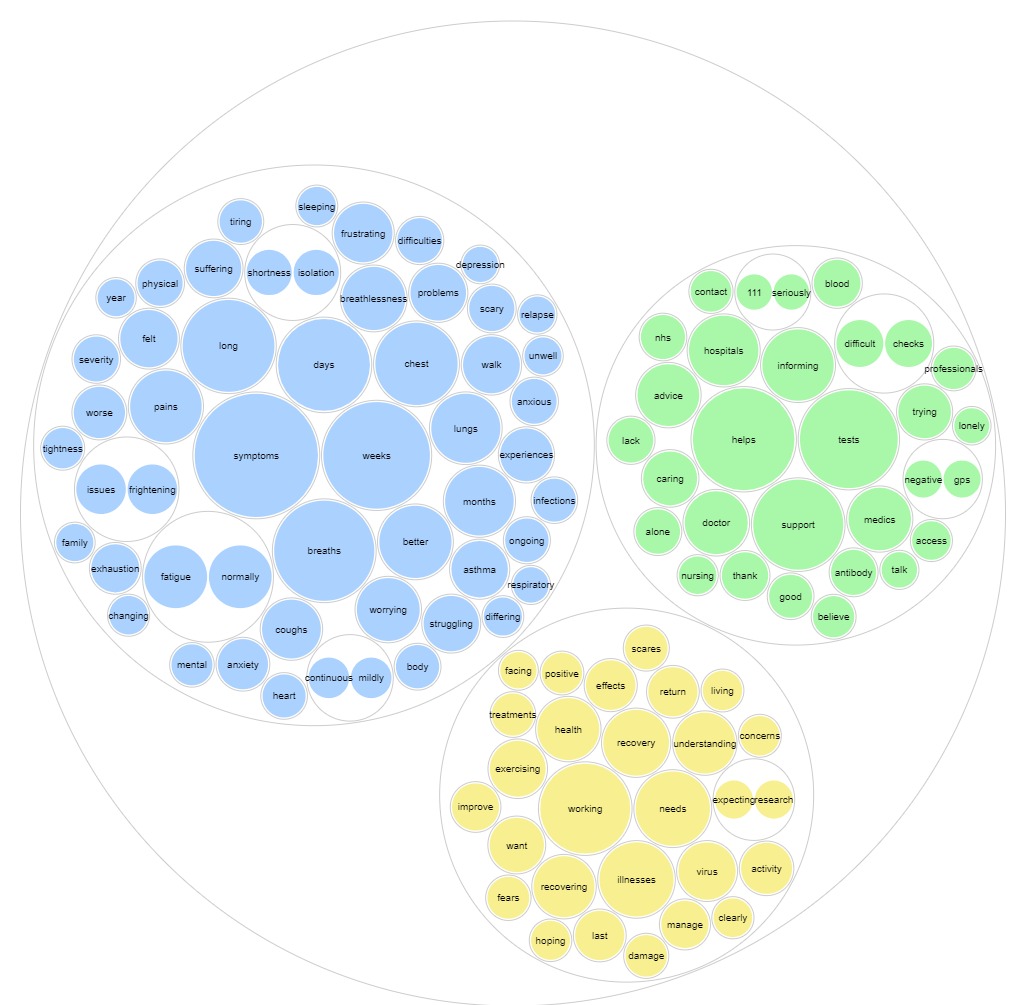
