## Supplement 2 for "Patient symptoms and experience following COVID-19: results from a UK wide survey"

| **Symptoms n** (%) | **Whole cohort**  **n=3023** | **Self-treatment at home cohort n=2644** | **Hospitalised patients n=376** | **p-value** |
| --- | --- | --- | --- | --- |
| Breathing problems - for example chest tightness, or struggling to breathe while resting or being active | 3027 (92.0) | 2646 (92.1) | 381 (91.4) | 0.61 (X) |
| Changes in mood, or anxiety or depression | 1417 (43.1) | 1214 (42.3) | 176 (42.2) | 0.70 (X) |
| Cough | 1392 (42.3) | 1207(42.0) | 185 (44.4) | 0.36 (X) |
| Extreme tiredness (fatigue) or lack of energy | 2739 (83.3) | 2394 (83.3) | 345 (82.7) | 0.76(X) |
| Hair loss | 346 (10.5) | 304 (10.6) | 42 (10.1) | 0.75 (X) |
| Loss of appetite or weight loss | 755 (22.9) | 657 (22.9) | 98 (23.5) | 0.77 (X) |
| Loss of taste or smell (anosmia) | 936 (28.4) | 808 (28.1) | 128 (31.9) | 0.28 (X) |
| Muscle weakness or joint stiffness | 1662 (50.5) | 1449 (50.4) | 213 (51.1) | 0.81 (X) |
| Nightmares or flashbacks | 432 (13.1) | 379 (13.2) | 53 (12.7) | 0.79 (X) |
| Problems with mental abilities; e.g. not being able to remember some events, think clearly and being forgetful | 1508 (45.8) | 1325 (46.1) | 183 (43.9) | 0.39 (X) |
| Sleep problems | 1520 (46.2) | 1341 (46.7) | 179 (42.9) | 0.15(X) |
| Symptoms of post-traumatic stress disorder (PTSD) - for example, feelings of isolation, irritability and guilt | 578 (17.6) | 503 (17.5) | 75 (18.0) | 0.81 (X) |
| **Average length of time between onset of symptoms and completing survey (days)** | 105.9±53.0 | 105.7±55.7 | 106.8±61.5 | 0.69 (¥) |

**Table E1:** *symptoms in a) whole cohort; b) Hospitalised patients and average time between onset of symptoms and completion of survey.*

*Data are presented as n (%) or mean ±. P values of <0.05 are taken to indicate statistical significance and are marked in bold. X= Chi square test used, ¥= t=test used. Where numbers do not total 100% missing values are due to questions unanswered by participant*

| **Effect of pre-existing conditions on long-COVID symptoms**   a) **Breathlessness in those with or without pre-existing lung disease** | | | | |
| --- | --- | --- | --- | --- |
| Count | | | | |
|  | | breathlessness | | Total |
|  |  | no breathing problems | breathing problems |  |
| lung disease | no pre-existing lung disease | 388 | 1993 | 2381 |
|  | pre-existing lung disease | 105 | 804 | 909 |
| Total | | 493 | 2797 | 3290 |

| **Chi-Square Tests** | | | | | |
| --- | --- | --- | --- | --- | --- |
|  | Value | df | Asymptotic Significance (2-sided) | Exact Sig. (2-sided) | Exact Sig. (1-sided) |
| Pearson Chi-Square | 11.624^a^ | 1 | .001 |  |  |
| Continuity Correction^b^ | 11.255 | 1 | .001 |  |  |
| Likelihood Ratio | 12.153 | 1 | .000 |  |  |
| Fisher's Exact Test |  |  |  | .001 | .000 |
| Linear-by-Linear Association | 11.621 | 1 | .001 |  |  |
| N of Valid Cases | 3290 |  |  |  |  |
| a. 0 cells (0.0%) have expected count less than 5. The minimum expected count is 136.21. | | | | | |
| b. Computed only for a 2x2 table | | | | | |

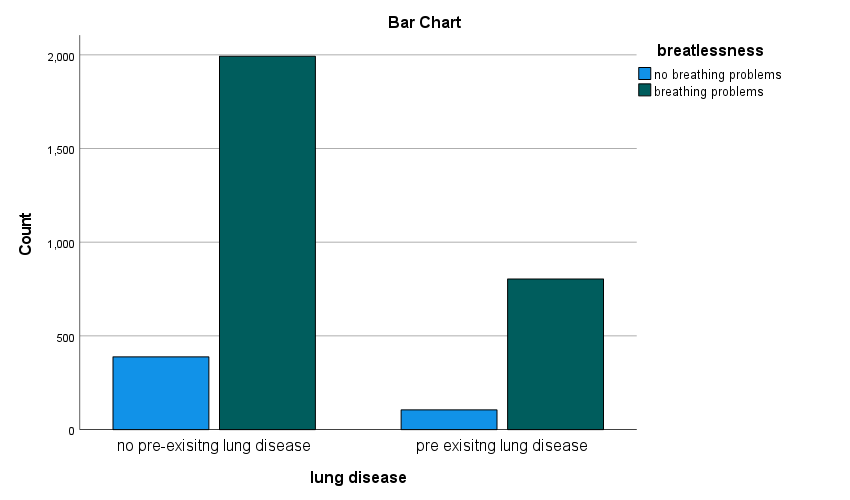

| b) **Mood disorder in those with or without pre-existing mental health diagnoses.** | | | | |
| --- | --- | --- | --- | --- |
| Count | | | | |
|  | | mood changes | | Total |
|  |  | no mood changes | mood changes |  |
| mood disorder | no pre-existing mental health disorder | 2163 | 625 | 2788 |
|  | pre-existing mental health diagnosis | 378 | 124 | 502 |
| Total | | 2541 | 749 | 3290 |

| **Chi-Square Tests** | | | | | | | | | |
| --- | --- | --- | --- | --- | --- | --- | --- | --- | --- |
|  | Value | df | Asymptotic Significance (2-sided) | | | Exact Sig. (2-sided) | | Exact Sig. (1-sided) | |
| Pearson Chi-Square | 1.262^a^ | | | 1 | .261 | |  | |  |
| Continuity Correction^b^ | 1.135 | | | 1 | .287 | |  | |  |
| Likelihood Ratio | 1.242 | | | 1 | .265 | |  | |  |
| Fisher's Exact Test |  | | |  |  | | .272 | | .144 |
| Linear-by-Linear Association | 1.261 | | | 1 | .261 | |  | |  |
| N of Valid Cases | 3290 | | |  |  | |  | |  |
| a. 0 cells (0.0%) have expected count less than 5. The minimum expected count is 114.29. | | | | | | | | | |
| b. Computed only for a 2x2 table | | | | | | | | | |

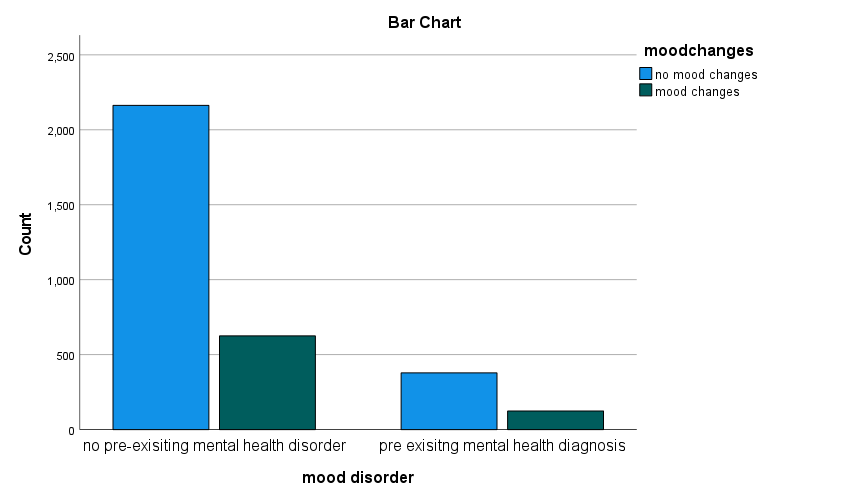

**Table E2:** Length of stay of hospitalised patient, ventilatory support and quality of communication/information received

|  | **Hospitalised patients n=417** |
| --- | --- |
| **How long were you in hospital for?**  Less than 1 week  1 to 2 weeks  2 to 4 weeks  More than 4 weeks | 238 (57.1%)  106 (25.4%)  51 (12.2%)  22 (5.3%) |
| **Were you able to breathe by yourself throughout your treatment for symptoms of COVID-19?**  Yes - I was able to breathe by myself without any additional support (apart from any medication that I normally take)  Yes - I was able to breathe by myself with some support (e.g. face mask or tube under your nose that delivers oxygen, CPAP machine)  No - I stopped being able to breathe by myself and needed mechanical support (e.g. ventilator tube that is put into your airways to help you breathe, ECMO life support where blood is pumped through a machine that acts as your lungs) | 140 (33.6%)  210 (50.4%)  44 (10.6%) |
| **How long were you in ITU for?**  Less than 3 days  4 to 7 days  7 to 14 days  More than 14 days | **n=102**  16 (15.7%)  33 (32.4%)  32 (31.4%)  20 (19.6%) |
| **Patient reported quality of communication/information received during hospital stay**  How clear was the information being communicated?  How useful was the information to you?  How timely was the communication?  How empathetic was the communication? | 5.6±2.9  5.3±3.0  5.4±3.0  6.4±3.1 |
| **Did you get a rehabilitation plan when you left hospital?** | Yes= 29 (7.0%)  No= 386 (92.6%) |

*Data are presented as n (%) or mean ±. P values of <0.05 are taken to indicate statistical significance and are marked in bold.* *Where numbers do not total 100% missing values are due to questions unanswered by participant.*

**Prevalence of breathlessness in ventilated and non-ventilated patients**

| **Ventilation * Breathlessness Crosstabulation** | | | | |
| --- | --- | --- | --- | --- |
| Count | | | | |
|  | | Breathlessness | | Total |
|  |  | no breathing problems | breathing problems |  |
| Ventilation | not ventilated | 26 | 324 | 350 |
|  | ventilated | 6 | 38 | 44 |
| Total | | 32 | 362 | 394 |

| **Chi-Square Tests** | | | | | |
| --- | --- | --- | --- | --- | --- |
|  | Value | df | Asymptotic Significance (2-sided) | Exact Sig. (2-sided) | Exact Sig. (1-sided) |
| Pearson Chi-Square | 2.019^a^ | 1 | .155 |  |  |
| Continuity Correction^b^ | 1.272 | 1 | .259 |  |  |
| Likelihood Ratio | 1.746 | 1 | .186 |  |  |
| Fisher's Exact Test |  |  |  | .150 | .131 |
| Linear-by-Linear Association | 2.013 | 1 | .156 |  |  |
| N of Valid Cases | 394 |  |  |  |  |
| a. 1 cells (25.0%) have expected count less than 5. The minimum expected count is 3.57. | | | | | |
| b. Computed only for a 2x2 table | | | | | |

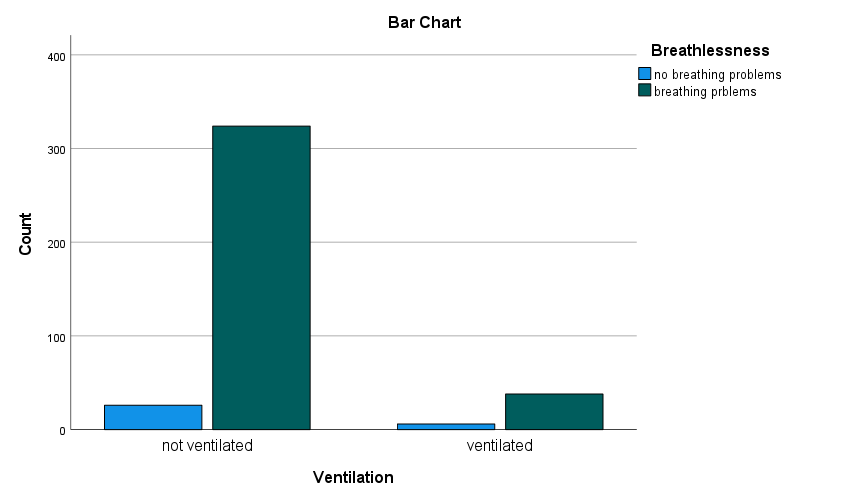

**Table E3:** Experience of care received during and after COVID-19 and ability to cope in a) whole cohort; b) hospitalised patients

|  | **Whole cohort**  **n=3290** | **Hospitalised patients n=417** | **P-value** |
| --- | --- | --- | --- |
| **Have you spoken to Nurse/GP?**  Yes  No | 2357 (71.6)  901 (27.4) | 282(67.6)  108 (25.9) | 0.21(X) |
| **Did your GP or nurse tell you about *things you can do* to help your recovery?**  Yes  No | 919 (27.9)  1428 (43.4) | 120 (28.8)  161 (38.6) | 0.16(X) |
| **Patient reported quality of communication/information received after COVID-19**  How clear was the information being communicated?  How useful was the information to you?  How timely was the communication?  How empathetic was the communication? | 5.6±2.7  4.7±2.8  4.9±3.0  6.1±3.1 | 6.0±2.7  5.4±2.8  5.3±3.0  6.6±3.0 | **0.002(¥)**  **P<0.001(¥)**  **0.025(¥)**  **0.004(¥)** |
| **How important are each of the following for you?**  General information on difficulties after COVID-19  Advice from healthcare professionals on managing difficulties after COVID-19  The latest medical research on difficulties after COVID-19  Reading about other people that have difficulties after COVID-19 and their experiences  Talking to other people with difficulties after COVID-19 and sharing experiences | 8.9±1.7  9.0±1.7  8.9±1.7  8.4±2.0  7.4±2.6 | 9.1±1.8  9.1±1.8  8.9±1.9  8.4±2.1  7.8±2.4 | 0.88(¥)  0.22(¥)  0.44(¥)  0.35(¥)  0.18(¥) |
| **Do you receive any practical help with your difficulties after COVID-19 on a regular basis?**  Yes, from someone living in my household  Yes, from someone living in another household  No, I don't need help  No, I can't get help | 1719 (52.2)  441 (13.4)  850 (25.8)  268 (8.1) | 247 (59.2)  79 (18.9)  56 (13.4)  33 (7.9) | **P<0.001** |
| **How much do you feel you can cope with your life after COVID-19?**  Completely  Mostly  Partly  Very little  Not at all | 269 (8.2)  1206 (36.7)  1279 (38.9)  454 (13.8)  82 (2.5) | 25 (6.0)  160 (38.4)  162 (38.8)  63 (15.1)  12 (2.9) | 0.40 |

*Data are presented as n (%) or mean ±. P values of <0.05 are taken to indicate statistical significance and are marked in bold. X= Chi square test used, ¥= t=test used. Where numbers do not total 100% missing values are due to questions unanswered by participant.*
